# Impacts of worldwide individual non-pharmaceutical interventions on COVID-19 transmission across waves and space

**DOI:** 10.1101/2021.03.31.21254702

**Authors:** Yong Ge, Wen-Bin Zhang, Haiyan Liu, Corrine W Ruktanonchai, Maogui Hu, Xilin Wu, Yongze Song, Nick W Ruktanonchai, Wei Yan, Eimear Cleary, Luzhao Feng, Zhongjie Li, Weizhong Yang, Mengxiao Liu, Andrew J Tatem, Jin-Feng Wang, Shengjie Lai

## Abstract

Governments worldwide have rapidly deployed non-pharmaceutical interventions (NPIs) to mitigate the COVID-19 pandemic. However, the effect of these individual NPI measures across space and time has yet to be sufficiently assessed, especially with the increase of policy fatigue and the urge for NPI relaxation in the vaccination era. Using the decay ratio in the suppression of COVID-19 infections, we investigated the changing performance of different NPIs across waves from global and regional levels (in 133 countries) to national and subnational (in the United States of America [USA]) scales before the implementation of mass vaccination. The synergistic effectiveness of all NPIs for reducing COVID-19 infections declined along waves, from 95.4% in the first wave to 56.0% in the third wave recently at the global level and similarly from 83.3% to 58.7% at the USA national level, while it had fluctuating performance across waves on regional and subnational scales. Regardless of geographical scale, gathering restrictions and facial coverings played significant roles in epidemic mitigation before the vaccine rollout. Our findings have important implications for continued tailoring and implementation of NPI strategies, together with vaccination, to mitigate future COVID-19 waves, caused by new variants, and other emerging respiratory infectious diseases.

## Main

The COVID-19 pandemic has caused significant disruption to daily lives, causing over 184 million confirmed cases and 4 million deaths as of 4 July 2021[1]. Non-pharmaceutical interventions (NPIs) have been deployed across the World to curb the pandemic[2]. With the rollout of COVID-19 vaccines using different dosing and population targeting strategies[3], robust vaccination programs would enable the relaxation of NPIs[4, 5]. However, given the delays in vaccine production and the inequality of vaccine allocations[6] as well as the emergence of novel variants[7, 8], NPIs should be maintained to avoid further resurgences before herd immunity can be achieved[4, 9, 10].

The impact of NPI policies might be dynamic, determined by a variety of factors such as policy fatigue and population immunity. First, because of variations in the government’s execution of NPIs and the degree of people’s inclination to comply, the same NPI may work differently in different regions. Second, over a lengthy period adopting NPIs, people tend to experience psychological tiredness, reducing the effectiveness of NPIs[11]. Third, changes in mutant viruses, vaccination rates, and immunity acquired from infections may have an impact on the efficacy of individual NPIs[12]. Given that there is a long way to go before herd immunity for COVID-19[13] is achieved, understanding the role of different NPIs in reducing COVID-19 transmission before vaccine rollouts is critical for tailoring effective NPI strategies for future COVID-19 waves and other epidemics caused by respiratory infections.

The effectiveness of NPIs on pandemic mitigation had been demonstrated by previous studies that mostly focused on the first wave of the pandemic[5, 14-18], with limited analysis of subsequent waves and multi-scale research[19]. The implementation of NPIs in the first wave had, to some degree, changed human knowledge and perceptions, behaviours and responses to mitigate the outbreaks[20-24]. The enduring importance of NPI responses to COVID-19 has also been highlighted[25]. Though policy fatigue has been proposed and confirmed in the implementation of NPIs[26], whether, and to what extent, NPI effectiveness decreases with fatigue in subsequent waves remains unclear. Additionally, the effects of NPIs may vary across countries, nations and subnational regions with different geographical characteristics, such as health capacity, residential population density, aging ratio, humidity and air temperature[27, 28]. The potential differences in NPI effectiveness across multi-geographical levels are rarely discussed in existing analyses[17].

In this study we estimated the effects of several individual NPIs as well as their combinations by identifying their contributions to the decay ratio of COVID-19 infections across waves at different geographical levels before the onset of any vaccination program. We used a national database, covering 133 countries, territories and areas, to estimate NPI efficacy at both global and regional scales, and a subnational database, covering 51 states of the United States of America, to evaluate national and subnational NPI efficacy in the USA. Both databases were publicly available with comparable outcomes, covering epidemiological[29], intervention policy[30], environmental and demographic data from the earliest available dates to 22 June 2021. The deployment time and intensity of nine NPIs, including school closures, workplace closures, public events closures, gathering restrictions, stay-at-home orders, internal movement restrictions, public transport closures, international travel restrictions, and facial coverings, were considered in the data processing.

## Defining waves and groups

### Waves

The inequality in pandemic development across the world has led some countries to confront more than one COVID-19 wave[31, 32]. To identify potential variation in effects of NPIs across waves, we divided the epidemic waves in each country/state based on the smoothed daily reported cases. An epidemic wave constituted a period of three or more consecutive weeks in each country/state, when the daily numbers of cases within this period all exceeded 5% of the maximum daily number of cases in 2020 in corresponding countries/states. The first and last days of these defined time periods were the start and end of the corresponding wave, respectively. Noting that the first wave of the pandemic in most countries/states began with low-level community transmission caused by imported cases, we adjusted the start date of the first wave. It was set to the day when the number of daily new cases exceeded 10 cases for countries where the maximum number of daily new cases in the first wave were no more than 300 cases. Otherwise, the start date was set to the day when the number of daily new cases exceeded 20 cases. The details and full lists of waves by country/state can be found in SI. Up to now, no more than three waves of epidemics have been detected before the implementation of mass vaccination in most countries/states.

### Regional stratification

The reported COVID-19 morbidity and mortality showed obvious spatial stratified heterogeneity among different countries/states, based on the released epidemiological data. A spatial variance analysis method known as a geographical detector model was used to divide the study countries/states into different groups, according to each country/state’s overall morbidity and mortality during the whole research period. The principle of the stratification method is minimizing variance within groups while maximizing variance between groups[33, 34]. Spatial proximity was also considered within groups because nearby countries/states were prone to have similar policies, intervention methods, as well as environmental conditions. We investigated the spatial variation in NPIs effectiveness by dividing 133 countries into four groups at the regional level and 51 states into three groups at the subnational level based on their COVID-19 morbidity and mortality together with geographical proximity (SI Fig. C2). Thresholds of 1,800 per 100,000 persons for morbidity and 40 per 100,000 persons for mortality were determined by q-statistic index in the geographical detector model to select countries with both high morbidity and high mortality. Considering the geographical proximity between countries, Asian countries and African countries were stratified into two separate groups. A full list of countries in each group and the corresponding time frame of different waves of COVID-19 can be found in SI Table C1 – C4, C6 – C8.

**Fig. 1.**
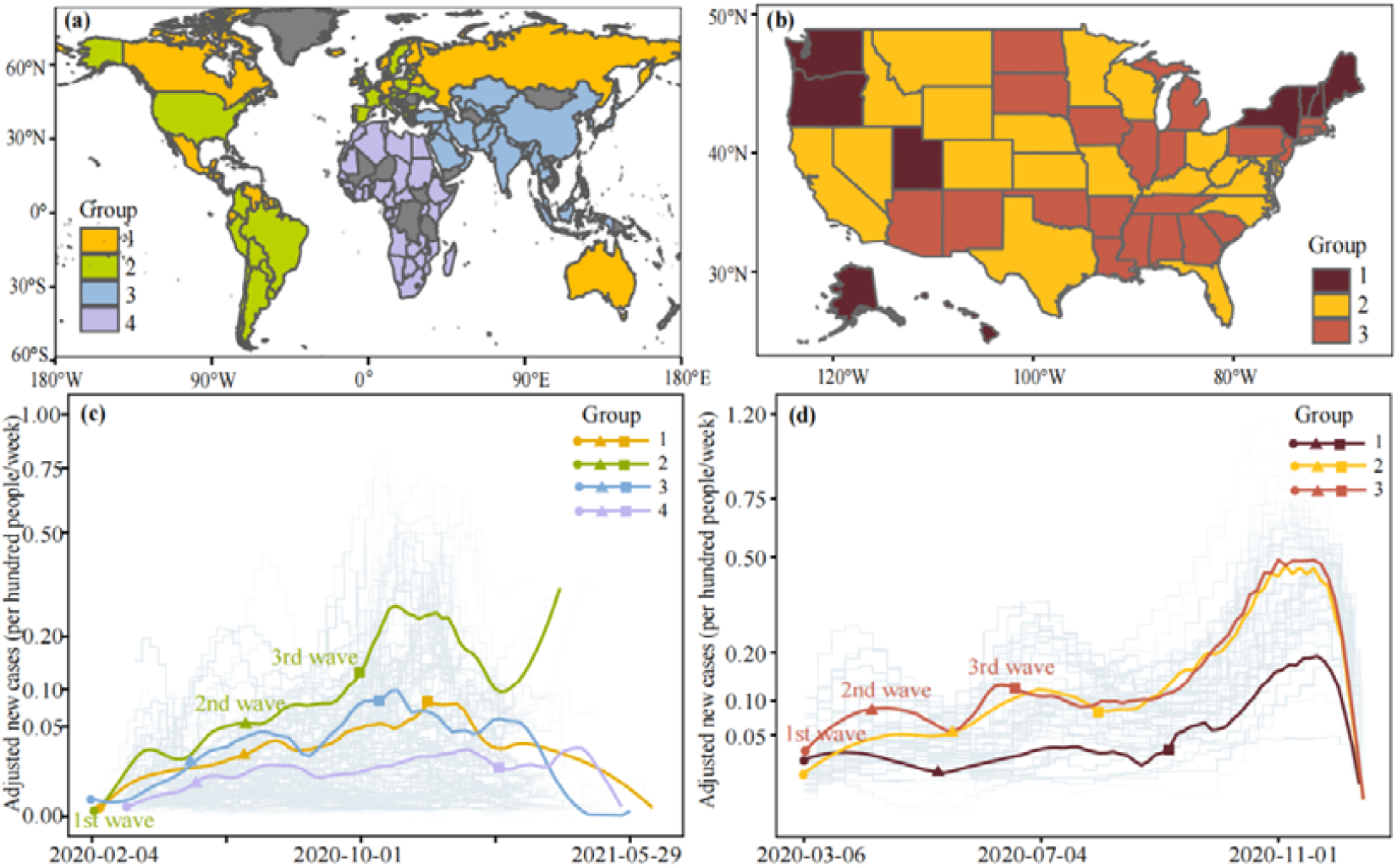
Data context. The different groups in (a) and (b) were determined by pandemic parameters and geographic proximity (see SI for more information). (c) -(d) The pandemic trajectories of weekly reporting cases for each country/state (background polylines) and group (solid curves). The solid curves for each group are mean weekly cases across countries/states in that group. Starting points (represented by different marks) of different waves in each group are generally illustrated by their mean starting dates which were quantitatively defined in this study.

### Spatiotemporal Bayesian inference model-based NPI efficacy evaluation

A Bayesian inference model[15, 20] was built to disentangle the individual effects of NPIs from the empirical changes of weekly growth rates. We measured relative contributions of NPIs on the observed decay ratio of COVID-19 infections (denoted as %Δ*ω*_*t*_) using the existence and intensity change of these interventions. The decay ratio was defined as a percentage of reduction in the baseline growth rate by the instantaneous growth rate. The instantaneous growth rate of transmission at each point of time was calculated as the current weekly number of new infections over the infections in the previous week. In addition to interventions, there were many other factors (e.g., the transmissibility of new variants and the variation of case diagnosis and reporting) that might affect the growth rate of COVID-19 transmission over time. Therefore, the baseline growth rates in different waves and countries were assumed as the mean of the top three highest instantaneous growth rates in the corresponding wave and country.

We used the decay ratio directly derived from the reported case data, rather than the effective reproduction number (*Rt*)[15, 17], to avoid introducing the uncertainty of estimating *Rt* over time[35]. The growth rate reflects how quickly the numbers of infections are changing day by day. It is an approximation of the percentage change in the number of infections each day. If the growth rate is greater than 1, then the epidemic is growing, otherwise it is shrinking. The growth rate provides us with information on the size and speed of change, whereas the *Rt* value only gives us information on the direction of change. Different diseases with the same *Rt* can generate epidemics that grow at very different speeds. Therefore, the growth rate may better reflect the pandemic change attributed to variants of concern. However, a relatively small number of cases in the States of the USA may correspond to greater uncertainty in estimated growth rates leading to a less robust evaluation of NPI efficacy as there will be a wider range given for growth rate and frequent changes in the estimates. This indicates that our model will be superior on larger scales for both *Rt* and the growth rate. However, estimation of the growth rate requires fewer assumptions about the disease than *Rt*.

We modelled NPI effects over time without assuming a functional relationship between effectiveness over time, which allows for variable community responses to the variation of each intervention in separate waves. The effect of each individual NPI across countries was characterized by the same coefficient, whereas the variation in efficacy for different countries was represented by adding Gaussian noise to our model for each single estimation. Then, the observed decay ratios for each country in each week were decomposed into NPI effectiveness according to the corresponding intervention timing and intensity, controlled for confounding using covariates which included health capacity, residential population density, aging ratio, humidity, and air temperature. All estimations were performed using Markov chain Monte Carlo (MCMC) methods. The reliability of our model was assessed by the cross-validation for overall intervention effects. Sensitivity analyses were also performed to assess model robustness in terms of our assumptions. More details on models and covariates can be found in Methods and SI.

### Global effects of individual NPIs across waves

We estimated all mitigation strategies of immediate interest before the start of mass vaccination implementation, where the four NPIs with the highest impacts (>30%) on transmission growth rate included school closures (median 36.8%, interquartile range [IQR] 27.0 -48.3%), international travel restrictions (36.0%, 26.3 -40.2%), facial coverings (33.6%, 27.0 -40.4%) and gathering restrictions (31.7%, 27.2-45.4%) (Fig. 2). The NPIs with moderate effects (25% -30%) included workplace closures (28.3%, 27.7-31.8%) and public transport closures (25.6%, 22.9 -35.9%), while movement restrictions had relatively limited impacts. The overall synergistic effectiveness of these NPIs reached 92.3% (IQR: 88.1-96.9%) and declined with the epidemic process of COVID-19, from 95.4% in the first wave to 56.0% in the third wave.

**Fig. 2.**
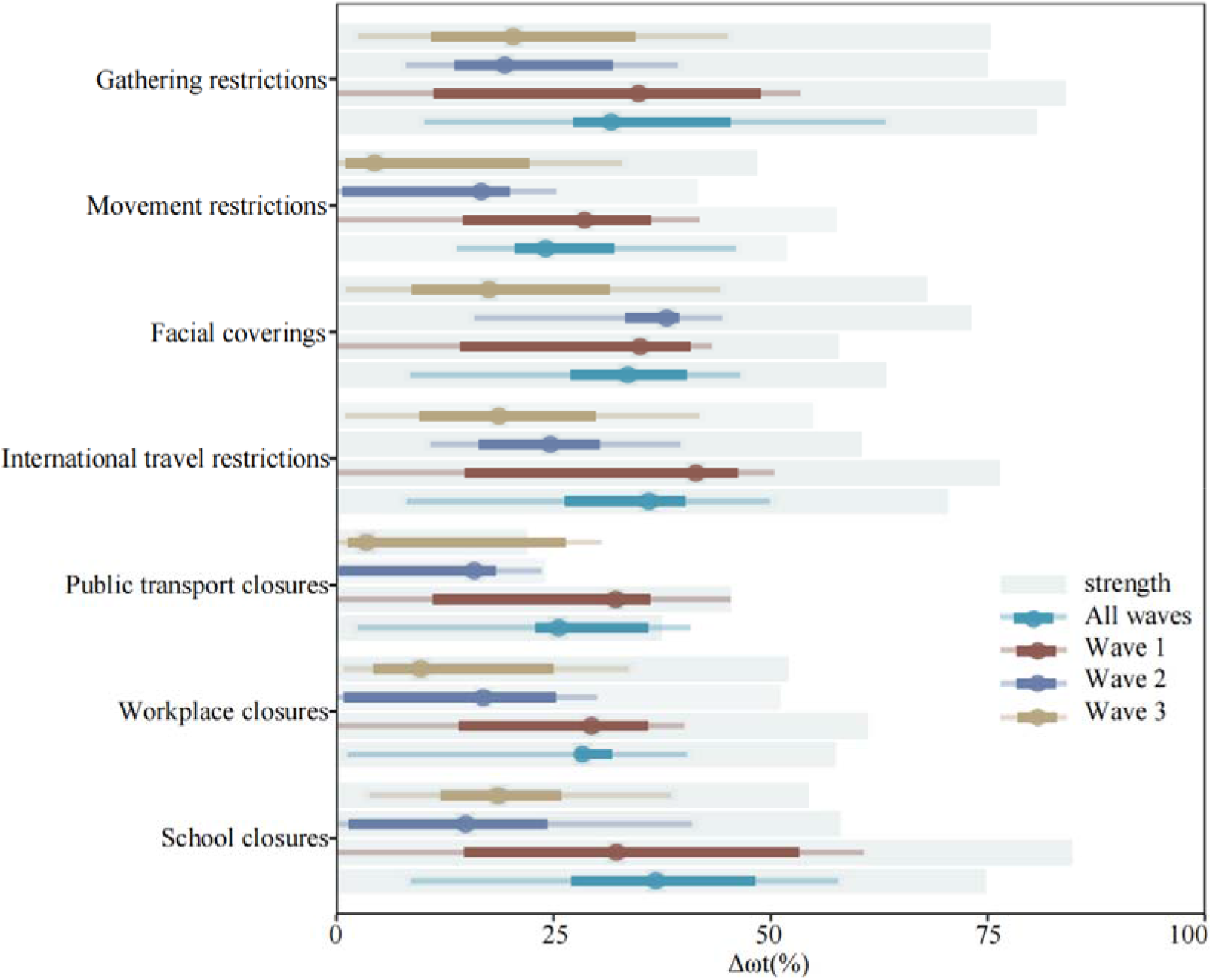
Effects of individual NPIs on reducing the transmission of COVID-19 across waves within our data context. The coefficients (*α*_*i*_) of NPIs parameters in different periods were calibrated by the default model setting with corresponding data contexts. The effect estimates were calculated by the coefficients of NPIs through 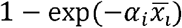, where 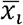 is the average strength of NPI implementation (represented by the background shadow). We rescaled the average strength by multiplying 100 to adapt the x-axis. The synergistic effectiveness of all NPIs (All waves: 92.3%, Wave 1: 95.4%, Wave 2: 79.9%, Wave 3: 56.0%) were nonlinear cumulative in terms of the individual effect by 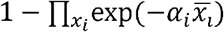. The effect over all waves represents the average performance of NPIs against COVID-19 in 133 countries (Fig. 1(a)) before their vaccination by 22 June 2021. Wave 1 refers to the average performance of NPIs against COVID-19 in the first wave of the 133 countries. The specific periods of the first wave in 133 countries are not fully consistent, meaning that the first wave does not refer to a particular time but a general period of the first outbreak. The second wave refers to the periods starting from the second outbreak. %Δ*ω*_*t*_ represents a decay ratio of the COVID-19 infection rate in each country. The 5th, 25th (Q1), 50th (median), 75th (Q3), and 95th percentiles of estimates of %Δ*ω*_*t*_ are presented to indicate details of the variations. The uncertainty intervals of NPI effectiveness refer to the variance over corresponding data contexts.

The effectiveness and dominance of NPIs varied across waves. In the first wave, the most effective NPI was international travel restrictions (median 41.4%, IQR 14.8-46.3%). Results also showed that the synergistic effectiveness of all NPIs exceeded 28% in the first wave. In the second wave, facial coverings became the NPI with the highest effect (38.0%, 33.2 -39.5%), while gathering restrictions became the most effective NPI in the third wave (20.4%, 10.9 -34.5%). In addition, the effects of workplace closures, public transport closures and movement restriction declined to 9.7 (IQR 4.2 -25.4%), 3.5% (1.3 -26.4%) and 4.41% (1.0 -22.2%) in the third wave, respectively.

### Regional NPIs impacts across waves by country group

Further, this study revealed that effects of individual non-pharmaceutical measures showed discernible spatial and temporal variations across countries and waves, when only the periods before vaccine rollouts were included for accurate evaluation of NPIs (Fig. 3).

**Fig. 3.**
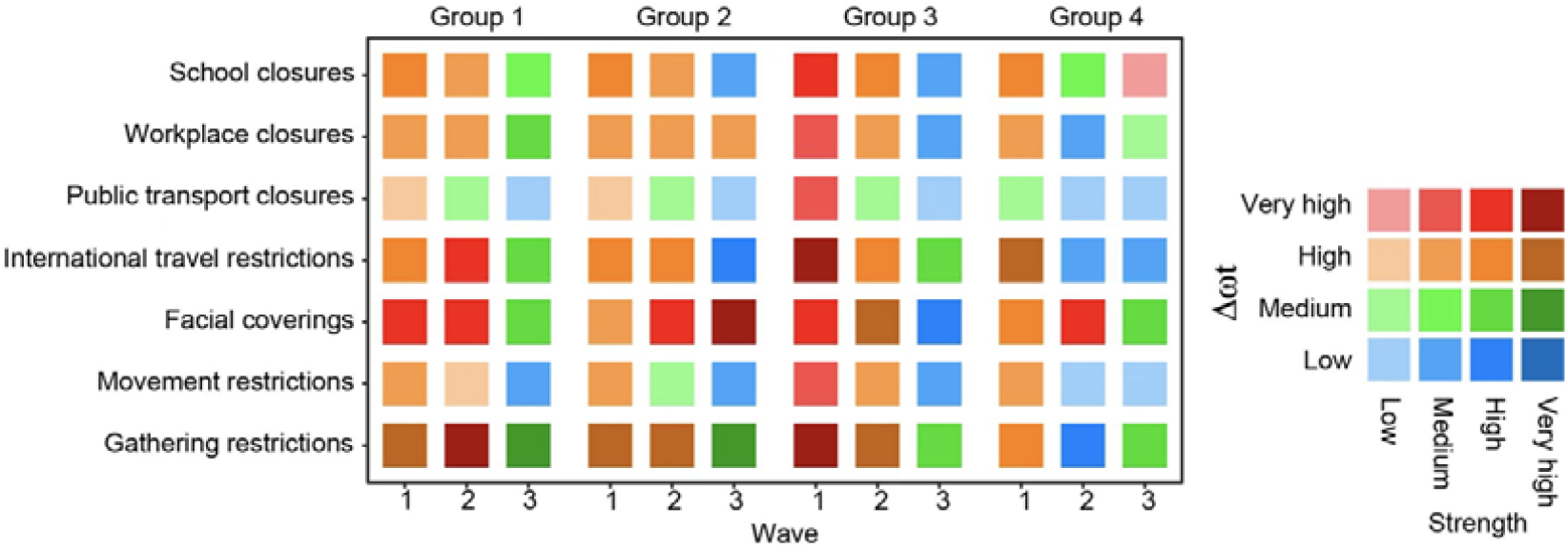
The cross-wave and cross-group effects of individual NPIs. Effects of individual NPIs on reducing the transmission of COVID-19 across waves and groups are illustrated by different colours associated with their average implementation strength. The dark colour indicates higher strength, while the NPIs effects increased from low level (blue) to high level (red). A full list of countries and the corresponding time frames of different waves for each group can be found in SI Table C2 – C5.

In the first wave, gathering restrictions in group 3 had the highest contribution (median 52.5%, IQR 30.5 -58.6%) to transmission reduction. We found all NPIs in Group 3, i.e., Asian countries, were generally more effective than other groups for the first wave, especially gathering restrictions (52.5%, 30.5 -58.6%), school closures (45.7%, 36.4 -54.6%) and facial coverings (43.7%, 35.4 -44.8%). In Group 4, all NPIs showed moderate effects (>20%), with the exception of public transport closure (17.4%, 11.8 -22.6%).

In the second wave, four NPIs in Group 1 had the highest effectiveness among four country groups. Facial coverings had an important role in reducing transmission for group 1 from the first to the second wave (%Δ*ω*_*t*_ >30%) and behaved similarly in the second Group’s second-to-third waves. Gathering restrictions and school closures exerted a relatively strong effect in both groups during the first two waves (>20%). International travel restrictions were one of the most effective NPI in affecting the pandemic transmission of Group 1 and made a notable contribution (median 33.52%, IQR 23.82 -41.91%) in the second wave. In Group 4, effects of NPIs were limited (<7%), except for facial coverings (41.3%, 37.6% -44.9%) and school closures (13.0%, 9.4% -17.8%).

After the first wave, as interventions were gradually relaxed, the effectiveness of most individual NPIs had declined by different degrees in different country groups. This decline was mainly observed in Group 2, i.e., the European, American, and Oceanian countries with relatively high morbidity and mortality, and Group 3, i.e., Asian countries. Countries in Group 1 and Group 2 were generally comprised of European, American, and Oceanian countries with relatively low and high morbidity and mortality, respectively. The highest effects of all NPIs in different waves were 97.9% for Group 3 in the first wave, 89.2% for Group 1 in the second wave, and 69.3% for Group 2 in the third wave. From the first to the third wave, effects of most individual NPIs were reduced, apart from facial coverings in Group 2 and school closures in Group 4, whose effects were increased. Workplace closures always played a mild role in controlling the spread of the virus in Group 2 for all waves but had limited effectiveness for Group 1 in the third wave.

In the third wave, effects of most NPIs had been critically reduced. The effectiveness of workplace closures (from 31.2% to 6.0% in median), public transport closures (from 33.2% to 5.8%) and movement restrictions (from 33.0% to 4.9%) declined from the first to the third wave. International travel restrictions were the only NPI which had stable effectiveness in all waves (>15%) for Group 3. Distinguishable from other groups, in Group 4, i.e., African countries, five out of seven NPIs had the lowest effectiveness in the second wave and then climbed in the third wave, resulting in the joint effect of all NPIs in the third wave surpassing that in the second one. Facial coverings (median 41.3%, IQR 37.6 -44.9%) of the second wave and school closures (30.8%, 21.8 -36.6%) of the third wave were the only NPIs in this group which surpassed other groups in suppressing infection.

### National and subnational effectiveness of NPIs in the United States

This study used the USA, as well as its states, as prisms to explain the potential spatio-temporal heterogeneity of NPI efficacy on a national and subnational scale, respectively. To keep in line with the larger-scale analysis, trajectories of the USA-states cases were divided into three waves using the wave division method developed in this study. Similar to results at the global scale, the overall synergistic effectiveness of NPIs showed a downward trend as the COVID-19 pandemic spread across the USA, from 83.27% (IQR 82.06-84.56%) in the first wave to 58.74% (57.01-60.33%) in the third wave, of which gathering restrictions and stay-at-home orders had the highest effect (>15%) on mitigating outbreaks. NPIs which had moderate impact were facial coverings and school closures (>10%), while public transport closures and workplace closures had limited effectiveness (<5%).

**Fig. 4.**
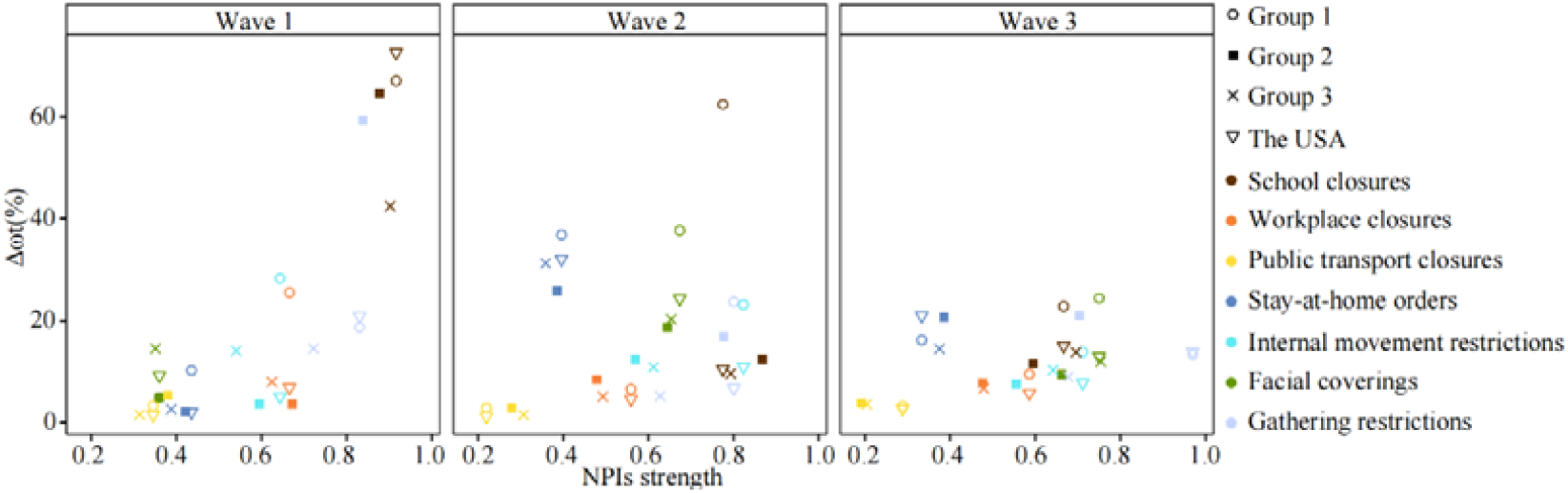
The individual efficacy estimates of the seven NPIs for the USA and its subnational regions. Different NPIs are represented by different colours. The groups as well as the USA are demonstrated by different symbols. Full lists of states name in each group as well as their defined wave periods can be found in SI Table C6-C8.

From a temporal perspective, the variability of NPIs was assessed through comparison of effects of NPIs across state groups. In the first wave, school closure was the primary NPI that was associated with a significant decline in incidence (>40%). The association between school closures and incidence increased with morbidity and mortality among groups. Regardless of school closures, the other six NPIs with lower effectiveness were internal movement restrictions (median 33.9%, IQR 28.3-43.5%), workplace closures (25.5%, 18.4-32.8%) and gathering restrictions (19.1%, 13.5-25.0%) in Group 1. In Group 2, gathering restrictions had the highest effectiveness (59.3%, 54.8-63.3%) beneath school closures (64.5%, 58.8% -69.4%), while facial coverings (14.5%, 12.7-16.2%) worked in parallel with internal movement restrictions (14.1%, 10.8-17.3%) and gathering restrictions (14.5%, 11.3-17.9%) in Group 3. In the second wave, school closures still had the highest effectiveness in controlling transmission in Group 1. Facial coverings significantly enhanced the effectiveness in Group 1, from 4.9% (3.2-7.2%) in the first wave to 37.7% (32.3-42.5%) in the second wave. In addition, stay-at-home orders had similar contributions, from 2.2% (1.4-3.3%) to 25.8% (20.3-31.0%) in Group 2 and 2.6% (1.7-3.9%) to 31.2% (25.6-36.3%) in Group 3. In the third wave, primary NPIs included facial coverings (24.4%, 16.2-32.8%) and school closures (22.8%, 15.8% -29.8%) in Group1; gathering restrictions (21.1%, 15.3-26.7%) and stay-at-home orders (20.7%, 15.7% -25.4%) in Group; and stay-at-home orders (14.5%, 9.8-19.5%), school closures (13.8%, 8.9% -19.3%), facial coverings (12.0%, 7.8% -17.1%), internal movement restrictions (10.3%, 6.6% -14.7%), and gathering restrictions (9.0%, 5.6% -13.5%) in Group 3.

## Discussion

Based on longitudinal public health interventions and socio-demographic datasets across COVID-19 waves, our study revealed that NPI measures played overwhelming roles in mitigating the pandemic, with varied effects across multi-spatial and temporal scales. Before the implementation of mass vaccination, the effectiveness of each individual NPI had been over 24.1% on the global level, 10.7% on the regional level, 1.0% on the USA national level, and 1.2% on the USA subnational level. Regardless of geographic scale and pandemic wave, the overall impact of integrated NPIs had been over 52.9% before the start of mass vaccination. Our results presented individual and synergistic NPI effectiveness in global, regional, national, and subnational scales, and this study was the first impact assessment that extended the research period from the beginning of the epidemic to before the vaccine rollout, to our knowledge. These findings are crucial for continued tailoring and implementation of NPI strategies to mitigate COVID-19 transmission among future waves (e.g., because of new variants of concern) or similar emerging infectious diseases, such as pandemic influenza.

On a global level, the synergistic effectiveness of NPIs had been declining along the waves before the start of mass vaccination. Lockdown fatigue or pandemic-policy fatigue might be the main reason for this phenomenon. Previous studies have found pandemic-policy fatigue to be geographically widespread, based on self-reported behaviours from a million respondents[26, 36]. Reported adherence to high-cost and sensitising interventions, like movement restrictions, decreased, while reported adherence to low-cost and habituating interventions, like facial coverings, increased in 2020. In our findings, the global efficiency of movement restrictions declined from 28.56% (IQR 14.56-36.25%) in the first wave to 16.67% (0.65-20.00%) in the second wave, while that of facial coverings climbed from 34.98% (14.24-40.82%) to 38.03% (33.22-39.48%), which is in accordance with the emergence of pandemic-policy fatigue. Our results show that the synergistic impact of NPIs and even the effect of individual facial coverings in the recent third wave was less than that in the second, suggesting the fatigue might continue to increase before vaccine rollout. Fatigue might have spread from high-cost interventions to lost-cost ones. Therefore, emphasizing the importance of policy compliance may have to be put on the agenda by local policy makers in pandemic mitigation.

The synergistic effectiveness of NPIs was not found to decrease among all groups and across all waves, however. In Group 1, i.e., American, and Oceanian countries with relatively low morbidity and mortality, the integrated efficiency of NPIs peaked at the second wave. These countries, except for Russia that only had one wave in our analysis, were largely protected from the epidemic due to the international travel control in the first wave[37], and witnessed an increased infection rate from 0.52% in the first wave to 0.73% in the second wave. The increased infection rate tends to push local people to react to the outbreak and maintain vigilance, bringing a higher synergistic NPI effect in the second wave than that in the first one. In Group 2 and Group 3, i.e., American, and Oceanian countries with relatively high morbidity and mortality, and Asian countries, respectively, the synergistic impact of NPIs decreased with each wave. They both experienced the most violent epidemic attack in the first wave, and thereafter, a better understanding of the coronavirus reduced people’s anxiety about the epidemic. Due to economic reasons and people’s expectations of recovery, the implementation strength of NPIs declined from average of 0.67 in the first wave to 0.63 in the third wave in Group 2, and from 0.69 to 0.52 in Group 3, further reducing the effect of NPIs. For Group 4, African countries, though the infection rate declined along the waves, the efficiency of integrated NPIs in the third wave surpassed that in the second wave. It is worth noting that many Africa nations might lack reliable epidemic data because of their reluctance to acknowledge epidemic or inadequate testing due to poverty and conflict[38]. Under-reporting confirmed case numbers and under-updated policy data would bring great uncertainty to the analysis results. Our results prove that the groups have different epidemic development and epidemic prevention trajectories. Countries should consider their own epidemic trajectory when learning from other countries’ experience in epidemic prevention and control.

There is partial consistency in NPI effectiveness at multiple geographical scales. Firstly, gathering restrictions and facial coverings both played significant roles in epidemic mitigation at a global scale and the USA national scale. The significant effects of gathering restrictions may be attributed to transmission most commonly occurring through spread of the virus in droplets or aerosols among people in close contact[39]. Therefore, appropriate levels of gathering restrictions and facial coverings should be maintained in subsequent waves before herd immunity is achieved. Secondly, the effect of school closures in epidemic control was significant at both the global and national level, with substantial heterogeneity between waves and geographic areas at both a regional and subnational level. The strongest effect of school closure was observed during the first wave at both global and the USA national level, as well as Group 3 at the regional level and Group 1 at the USA subnational level. School closures included not only primary and second level education institutes, but also universities which may serve as a bridge population for family/community transmission of the coronavirus[40]. In areas where school closures have had a good effect on epidemic control, this NPI can still be used during the vaccine delivery stage. Thirdly, international travel restrictions had a stronger protective effect than movement restrictions at both a global and regional level, while movement restrictions, especially stay-at-home orders, reflected its protective role at the national and subnational scale. Countries that quickly implemented border controls might have reduced the seeding of COVID-19 between countries, but international travel restrictions cannot prevent local transmission at the community level in countries where the virus had already been introduced. The increased effect of movement restriction on infection transmission, contributed to by both stay-at-home orders and internal movement restrictions in our study, at national and subnational level proved that our model has a good explanatory power for the effect of NPIs on epidemic migration.

It should be noted that our research limited the study period from the start of case reporting to the start of vaccination in each country. Therefore, pandemic-policy fatigue which happened before the vaccination rollout might worsen in the vaccination era. However, studies have shown that vaccination alone is insufficient to contain the outbreak, even with the most optimistic assumption of 85% infection prevention of vaccines[25]. Especially for the poor and people living in areas with low resources [6]. Non-pharmaceutical interventions should therefore not be rapidly relaxed in the vaccination era, considering the emergence of new variants and the inequality of vaccine delivery among countries[41].

We acknowledge that there are limitations in our analysis. First, data collected from public data sources may generate certain uncertainty. We did not make our own datasets from the sources but used publicly released ones from Johns Hopkins University, CDC, OxCGRT, United Nations, AHA, and so on. Data produced by different institutions may have differences in data due to subtle differences in statistical calibres and related regulations. Due to the huge amount of data and the relatively reliable data sources, in the absence of obvious inconsistencies in the data, we did not analyse the differences in data source statistics and uncertainty, which might bring uncertainties to the results. Second, the interactions among the seven NPIs were not considered in this study. There are differences in the way in which NPIs interact in each region. Considering the complex timeline of NPI interaction on a large scale, our study gives the individual effect of each NPI and the joint effect of all NPIs. The individual effect allows the comparison of NPIs effectiveness in each wave and geographical level, while the integrated effect, not linear addition of individual effects, presents the comprehensive effect of all NPI interactions. Third, we used grouped regional research instead of researching each country while the effects of NPIs might have some differences in different countries. However, the group results should be more robust than that from a single country because of a larger number of samples in each group considered. The effects of NPIs within each group were assumed following gamma distribution based on previous studies[15, 16]. The obtained different NPI effects reflected the overall effects in different waves and groups while varied in different countries.

Overall, the disclosure of epidemic, publicized responses allow us to estimate and compare the cross-wave effects of public health measures at global, regional, national, and subnational scales. Our work provides a quantitative basis and approach to explore historic spatio-temporal variation in the effectiveness of individual NPIs before the mass implementation of vaccination. The overall effectiveness of NPIs shows a downward trend between waves, possibly due to policy fatigue. Even though the synergistic efficiency of NPIs has been over 50% in recent waves before the vaccine rollout, the reducing effect over time deserves our vigilance, especially in areas lacking vaccines. Through the verification of multi-scale results, our study certified the effectiveness of gathering restrictions and facial coverings in the epidemic mitigation, which could be maintained in the following waves and mitigate pandemics caused by other emerging respiratory infectious diseases in the future.

## Methods

### Data sources and processing

#### Epidemiological data

The daily number of confirmed cases reported by country were obtained from the COVID-19 Data Repository by the Center for Systems Science and Engineering (CSSE) at Johns Hopkins University (JHU)[29]. While the state-level cases for the US were reported by the CDC[42]. The cases were recorded after infection-to-confirmation delay since the onset of their infection. The infection-to-confirmation delay was the sum of the incubation period and the delay from the onset of symptoms to confirmation. We first transformed the collected case data into their infection dates by an infection-to-confirmation delay following Negative binomial distribution [43-45]. To remove the influence of outliers and the fluctuation caused by the day-of-week effect, we smoothed daily case counts with the Gaussian kernel by calculating the rolling average using a Gaussian window with a standard deviation of 2 days, truncated at a maximum window of 15 days[46].

#### Intervention policy data

The non-pharmaceutical interventions studied in this work were: (1) school closures, (2) workplace closures, (3) gathering restrictions, (4) public transport closures, (5) movement restrictions, (6) international travel restrictions, and (7) facial coverings, collected and generated from the Oxford COVID-19 Government Response Tracker (OxCGRT)[30]. Five of the seven considered NPIs were directly used from OxCGRT in the global and regional analysis, i.e., school closures, workplace closures, public transport closures, international travel restrictions, and facial coverings. Gathering restrictions integrated public events cancellations and gathering restrictions, as the latter two NPIs documented in OxCGRT were highly collinear in terms of their timing and intensity of implementation across the 133 study countries. Similarly, movement restriction was produced by combining stay-at-home orders with internal movement restrictions. With respect to the national and subnational context, we replaced international travel control with internal movement restrictions and recovered movement restrictions to stay-at-home orders, due to the changing of collinearity and rare variation in international travel control across states of the US. The intensity of NPIs policies documented in OxCGRT was scaled into discrete values between 0 to 1 by dividing their maximum intensity, where 0 represented an absence of the NPI and 1 represented the corresponding maximum intensity. The intensity of school closures was further corrected as 1 during public and school holidays[47]. The intensities of integrated NPIs were calculated by the mean of their components’ intensities.

#### Environmental and demographic covariates

To control for country-specific confounders in the estimates of intervention effectiveness varied across countries, we also assembled population density, aging ratio, health capacity index, air temperature, and humidity for all these 133 study countries. Within each country, population density (per square kilometre) was the ratio of the total population over the corresponding built-up area in 2014[48]. The total and age-grouping population data in 2019 were obtained from the United Nations to calculate the aging ratio (> 65 year old) among populations[49]. Health capacity index was the arithmetic average of the five indices, including i) prevent, ii) detect, iii) respond, iv) enabling function, and v) operational readiness, developed to characterize the health security capacities in the context of the COVID-19 outbreak[50]. Air temperature and humidity were derived from the Global Land Data Assimilation System[51]. With respect to state-level data of the US, we used an alternative health capacity index, i.e., bed capacity, to capture the uneven distribution of hospital capacity relative to regional need, as well as substantial geographic variation in bed capacity per capita from 2012[52]. The measure was developed from three separate data sets: data from the American Hospital Association (AHA) when available, as well as data from the CMS Provider of Services file and CMS Cost Reports to fill in the gaps for hospitals that did not report their data to the AHA.

To further remove the day-of-week effect among case testing, diagnosis, and data reporting, all data used in this study were assembled and aggregated into a weekly dataset. The correlations between each two covariates were given to show their collinearity (SI, Fig. C1). The studied countries were selected by being documented in every dataset of epidemiological data, intervention policy data and environmental and demographic covariates. The details of data collection and processing are further provided in the Supplementary Information.

### Model description

#### Transmission dynamics

The evolution of the COVID-19 in a society can be characterized by 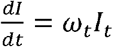, where *I* represents the new cases, and *ω*_*t*_ is the instantaneous growth rate. We adopted a general linear formula[15, 16, 53] linking NPIs to the empirical pandemic evolution. That is,

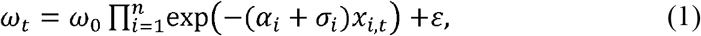

where *ω*_0_ represents the baseline growth rate without interventions, *α*_*i*_ is the coefficient of NPIs and control variables *x*_*i,t*_ on day *t*, and *ε* is the error term representing the uncertainty of decay ratio. In contrast to the control variables used to describe the country-specific difference in NPIs efficacy, we also introduced normal error term *σ*_*i*_ for *x*_*i*_ to capture the intrinsic variation of effectiveness across countries.The effect of NPIs set *X* in a period, such as the first wave of the pandemic, can be interpreted as a decay ratio in *ω*_0_ by computing 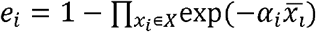 is the average strength of the NPI *x*_*i*_ during that period. The highest effect of NPIs set *X* where 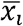 is 1, representing that the transmission is fully contained or interrupted by the set.

#### Computational environments of NPIs multiscale efficacy

We used the spatiotemporal Bayesian inference model to evaluate the effect coefficients in Eq. (1) based on the observed real-time COVID-19 growth rates, identifying the relative NPIs and vaccination effectiveness. We first evaluated the global effectiveness of NPIs for the whole data context. In addition to the overall NPIs effectiveness, we also evaluated NPIs regional effects in the first, second and third waves for each country group to show the potential large spatiotemporal diversity, respectively. Finally, national and subnational variation of NPIs efficacy in space and time was demonstrated by the case of the USA with 51 states. To exclude the vaccination impact on COVID-19 growth, we only used data before the onset of the vaccination project in all countries.

**Fig. 5.**
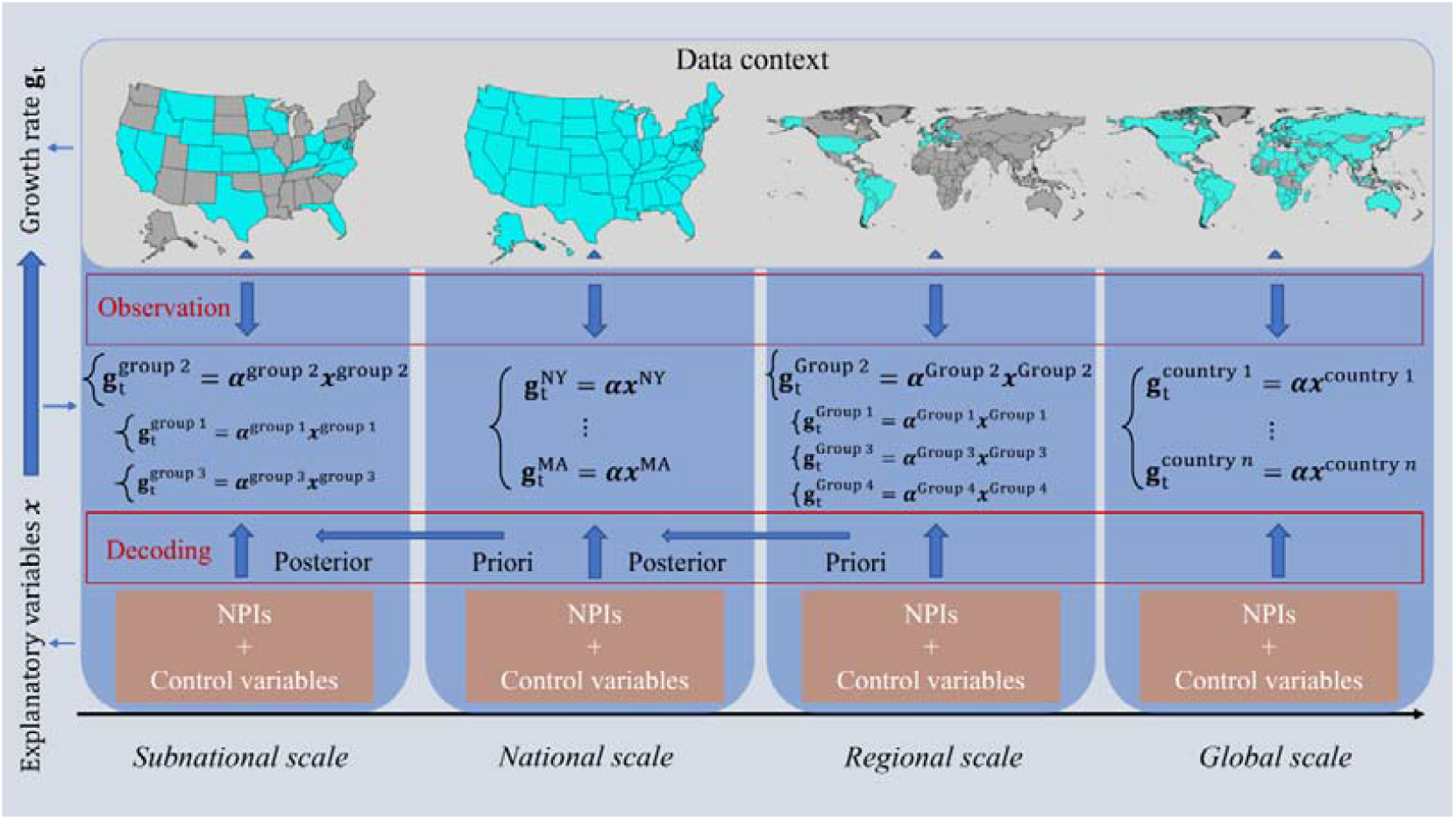
The conceptual framework. We used growth rate as the outcome variable to describe the trajectory of the pandemic. The empirical changes of growth rate were decoded into the effectiveness of both NPIs as well as the control variables.

We aimed to compare relative effectiveness of COVID-19 interventions across countries and across waves. Specifically, the effect of coefficients was assumed to follow a gamma distribution[16] in spite of NPIs and sociodemographic factors. Then, we placed 80% of their probability mass on positive effects for both NPIs and vaccination by left shifting their probability distributions with certain values[15].

While we put no information on the effectiveness of sociodemographic factors, i.e., placing 50% of their probability mass on positive effects. Under the circumstances, NPIs were more likely to contain the pandemic, while the state-variable effects were unknown. For the subnational analysis in the US, we used the national posterior of the effect coefficients estimated by the mix of all the states as a priori for each state to evaluate the state-level NPIs efficacy, alleviating the small sample effect on results robustness. The baseline growth rate (*w*_0_) was defined as the mean of the top three highest growth rates of the confirmed COVID-19 new cases in the corresponding wave. The instantaneous transmission interval (1/*ω*_*t*_) in the following weeks was assumed to have a gamma distribution also.

#### Model validation

The reliability of our models and corresponding results were evaluated by the leave-forty-countries-out cross validation. We first calibrated our model using 70% countries (93), randomly selected from 133 countries, to estimate the overall NPIs effects in both the first and second waves. Then, we derived the instantaneous growth rates through the estimated overall effects of NPIs for the remaining 30% countries (40) in terms of their implemented interventions. We used mean square error, ranging from 0 to infinite with 0 representing the perfect prediction ability, to assess the difference between the predicted instantaneous growth rates and the corresponding empirical instantaneous growth rates. We repeated this procedure 50 times, where the average mean square error was (median 1.4, interquartile range [IQR] 1.3 – 2.0). Further, we standardised the predicted and empirical instantaneous growth rates, respectively, within each country and then analysed all the data with one-way ANOVA.

#### Sensitivity analysis

The robustness of models and parameters used in the study was also assessed by a series of sensitivity analyses. The parameters to be assessed included: i) the probability mass of NPIs and vaccination on negative effectiveness and ii) the probability mass of sociodemographic factors on negative effectiveness. In this study, the default values for these parameters were 20% and 50%, respectively. The comparison of parameter impacts on estimates were listed in SI Table B1, representing three scenarios with smaller and larger default parameter settings. The differences of NPI effects among three waves were tested using a Wilcoxon signed-rank test, a non-parametric statistical hypothesis test for comparing NPIs effects between pairs of the three waves.

Using an R package, rstan[54], this model infers posterior distributions of each NPI effectiveness with the Markov chain Monte Carlo (MCMC) sampling algorithm. To analyse the extent to which modelling assumptions affect the results, our sensitivity analyses included epidemiological parameters, prior distributions, and the structural assumptions introduced above. MCMC convergence statistics are shown in SI Fig. B4.

## Data and code availability

All source code and data necessary for the replication of our results and figures are available at: https://github.com/wxl1379457192/NPIs_code

## Acknowledgments

This study was supported by the National Natural Science Foundation for Distinguished Young Scholars of China (No. 41725006), the Bill & Melinda Gates Foundation (INV-024911 and OPP1134076), the National Natural Science Foundation of China (81773498), and Beijing Natural Science Foundation (Grant No.7212128).

## Author contributions

YG, WBZ, HYL, WY, CWR and SJL conceived and designed the study, built the model, collected data, finalised the analysis, interpreted the findings, and wrote the manuscript. MGH, XLW, YZS and MXL collected data, interpreted the findings, and revised drafts of the manuscript. NWR, EC, JFW, LF, ZJL, WZY and AJT interpreted the findings, and commented on and revised drafts of the manuscript. All authors read and approved the final manuscript.

## Ethical approval

Ethical clearance for collecting and using secondary data in this study was granted by the institutional review board of the University of Southampton (No. 61865). All data were supplied and analysed in an anonymous format, without access to personal identifying information.

