## Supplementary material for "Impacts of worldwide individual non-pharmaceutical interventions on COVID-19 transmission across waves and space": SI

**Supplementary Information**

### A Supplementary method

#### **A.1 Detailed model description**

We employed a Susceptible-Infected-Removed (SIR) model[1] to describe the evolution of COVID-19 for each country. The infected populations at country $c$ on time $t$ were attributed to the number of COVID-19 cases on time $t-1$ by the following equation

$$I_{c,t}=\omega_{c,t-1}I_{c,t-1},$$

where $\omega_{c,t-1}$ was the instantaneous growth rate[2] of COVID-19, affected by the basic transmission rate (i.e. basic reproduction number), interventions, and the susceptible population ratio, while $I$ was the number of infections. The instantaneous growth rate was the inverse of the serial interval (the time between successive cases in a chain of transmission), which was commonly characterized by a gamma distribution[3]. Considering the huge susceptible populations at the early stage of the pandemic and vaccine rollouts, and no country has reached herd immunity[4], for the time being, we assumed that the basic growth rate in the pre-vaccination era with no interventions was approximately constant. However, the observed instantaneous growth rate ($I_{c,t}/I_{c,t-1}$), derived from the empirical daily confirmed cases, usually possessed a decreasing trend within each wave. For example, the first COVID-19 outbreak in China had a basic reproduction number of 3.54[5], but following the multipronged interventions, the reproduction number was reduced to 0.28 as of 8 March 2020. Nevertheless, most of the population in China are still susceptible to coronavirus by 31 March 2021, due to the sustained containment of the disease and the small proportion of Chinese vaccinated.

As previous works[6-8], we assumed that the decreasing transmission rate in each wave was contributed by interventions, especially NPIs in the pre-vaccination era. Then, we decomposed the variation in the empirical instantaneous growth rate in country $c$ into the variation in the timing and intensity of NPIs implemented during the corresponding period in country $c$. To this end, we modelled the efficacy of seven NPIs in country $c$ using the following formula,

$$\begin{aligned} \omega_{c,t-1}=\frac{I_{c,t}}{I_{c,t-1}}=\prod_{i=1}^{7} \exp\left( -\alpha_{c,i,t-1}x_{c,i,t-1} \right),\#\left( 1 \right) \end{aligned}$$

where $x_{c,i,t-1}$ is the state of NPI $i$ in country $c$ on time $t-1$ (see Fig. A3), and $\alpha_{c,i,t-1}$, the corresponding coefficient of NPI $i$, was used to measure the effect of each intervention. We assumed that $\alpha_{c,i,t-1}$ has a gamma distribution over time,

$$\alpha_{c,i,t-1}\sim Gamma(1/7, 1)-log(1.000382)/7.$$

We put this prior on $\alpha_{c,i,t-1}$ such that about 80% probability mass of NPI effectiveness on positive effect (see Fig. A2).

However, if there was rare variation in the timing and intensity of each NPI implementation, we cannot derive a reliable result from Eq. (1). We, thus, jointly estimated NPI effects through numerous countries by assuming $\alpha_{c,i,t-1}$ being constant across countries, and used Gaussian error $\sigma_{i}$ to capture the fluctuation between these countries. The social context-based variation of NPI effects across countries was further controlled by socio-demographic factors with the following equation,

$\begin{aligned} \omega_{c,t}=\omega_{c,0}\prod_{i=1}^{7} \exp\left( -\left( \alpha_{i,t}+\sigma_{i} \right)x_{c,i,t} \right)\prod_{j=1}^{5} \exp\left( -\left( \beta_{j,t}+\sigma_{j} \right)y_{c,j,t} \right), c\in\left\{ 1,\ldots,C \right\} \#\left( 2 \right) \end{aligned}$

where $\omega_{c,0}$ was the baseline growth rate in country $c$, and $\beta_{j,t}$, the coefficient of the socio-demographic factor $y_{c,j,t}$, was used as a proxy of the impact of this factor on the decay of the baseline growth rate. The correlation coefficients for the socio-demographic factors were also assumed to have a gamma distribution, but with 50% probability mass on their positive effects (see Fig. A2),

$$\beta_{c,j,t}\sim Gamma(1/5, 1)-log(1.09)/5.$$

However, the absolute values of instantaneous growth rate might vary heterogeneously across countries^8^. We added the baseline growth rate in Eq. (2) for country $c$ to make the NPI effects comparable across countries, by defining the NPI efficacy as a proportion of reduction from the baseline growth rate.

In the pre-vaccination era, to differentiate the performance of NPIs across waves and country groups, we first estimated the overall effect of NPIs covering the whole 133 countries as of the last days before their vaccination by 22 June 2021. Then, the relative NPI effects were evaluated in each country group and wave, respectively. To make Eq. (2) solvable, $\alpha_{i,t}$ were set to be constant for the practical estimation, where the variation in the NPI efficacy was captured by setting different data contexts in terms of space and time.

We estimated coefficients in Eq. (2) jointly for all countries within any particular above data context by an individual hierarchical model,

$$\frac{1}{\omega_{c,t-1}}\sim Gamma\left( \mu,\sigma\right),$$

$$\mu=\frac{1}{\omega_{c,0}\prod_{i=1}^{7} \exp\left( -\alpha_{i}x_{c,i,t-1} \right)\prod_{j=1}^{5} \exp\left( -\beta_{j}y_{c,j,t-1} \right)}.$$

The above model was fitted using an adaptive Hamiltonian Monte Carlo (HMC) sampler in Stan, a probabilistic programming language. We ran 5 chains for 300 iterations with 150 iterations of warmup and a thinning factor 1 to obtain 750 posterior samples. Posterior convergence was assessed using the Rhat statistic and by diagnosing divergent transitions of the HMC sampler.

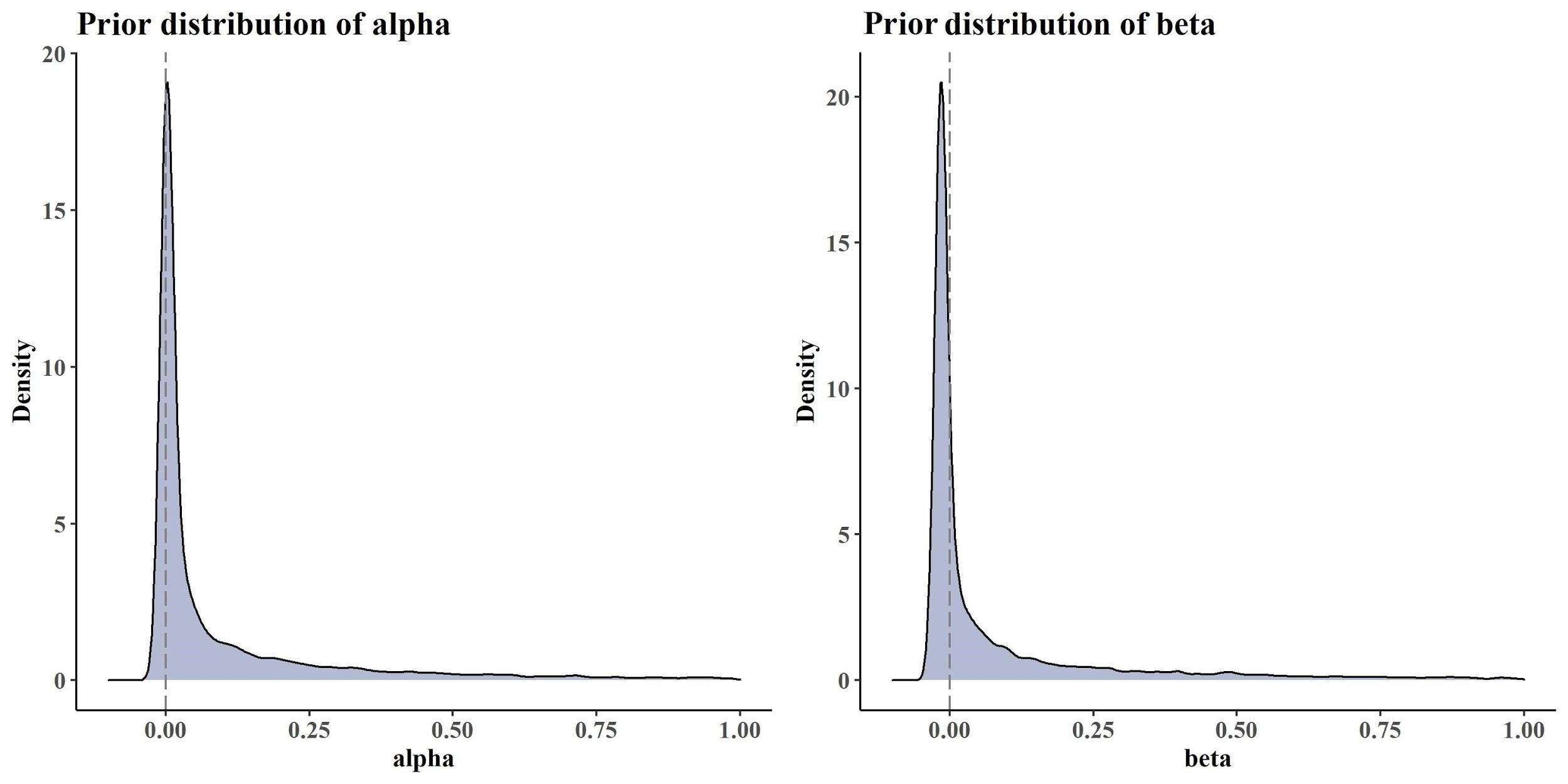

**Fig. A1.** The prior distribution of effect parameters of NPIs (left) and the correlation parameter of the socio-demographic factors (right).

#### **A.2 Details in data processing**

The datasets used in this study were all publicly available and detailed below. As different databases covering different countries, only countries documented in all these datasets were studied, including a total of 133 countries, territories and areas. Here, we detail how we assembled and processed each dataset into final integrated, experimental datasets for this study.

*Epidemiological data*

We collected daily confirmed COVID-19 case data for countries $c\in\left\{ 1,\ldots,133 \right\}$ from the earliest available dates to 25 March 2021. The daily confirmed cases used in our study were originally collected by the COVID-19 Data Repository by the Center for Systems Science and Engineering (CSSE) at Johns Hopkins University (JHU)[9], which can be obtained from the GitHub (<https://raw.githubusercontent.com/owid/covid-19-data/master/public/data/owid-covid-data.csv>). We processed epidemiological data country by country with the following steps: i) To account for the delay from infection to reporting, for each reported case, we generated a sample from a Negative binomial distribution where the mean was $N(10.92,\sigma=0.94)$ and dispersion was $N(5.41,\sigma=0.27)$ [10-12]. Then, we moved the case to its infection date by substrcting its reporting date with the sample (Fig. A2); ii) The daily numbers of cases were smoothed by calculating the rolling average using a Gaussian window with a standard deviation of 2 days, truncated at a maximum window size of 15 days[13]; iii) Defining periods of waves by our proposed quantitative method (see main text). A few country samples are illustrated in Fig. A3 to show the smooth procedure and waves.

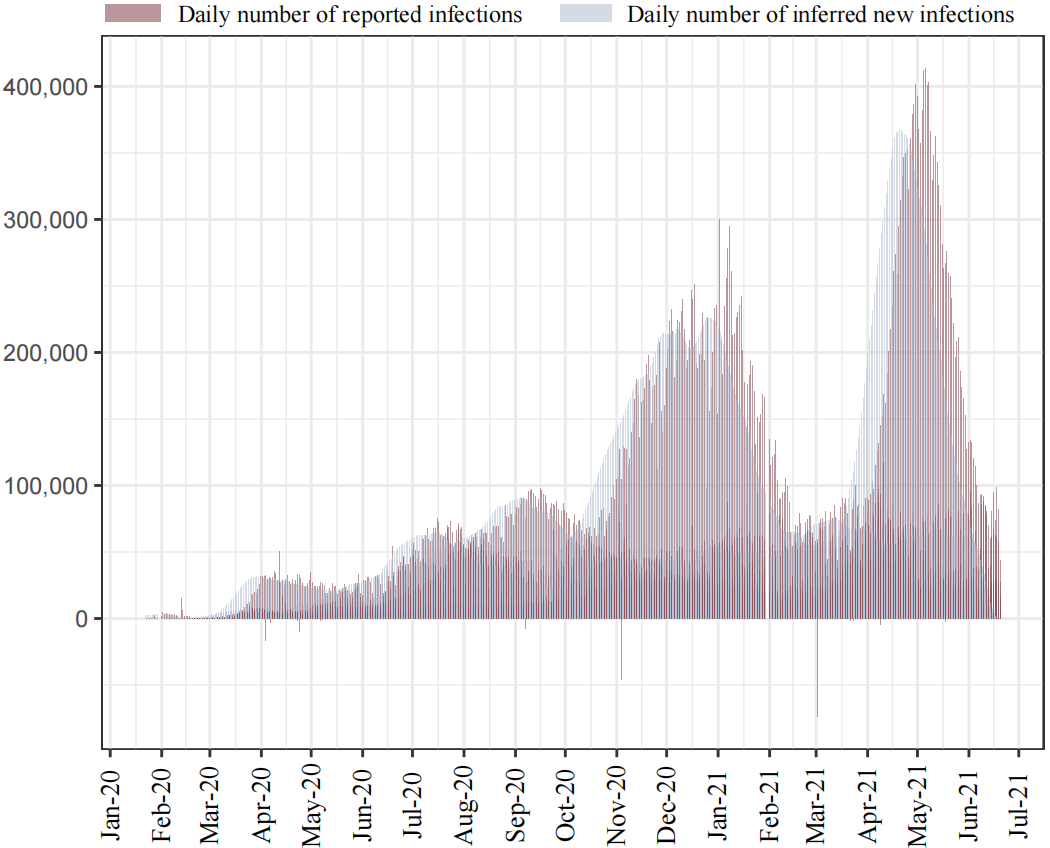

**Fig. A2.** The infection data transferred from the daily confirmed cases.

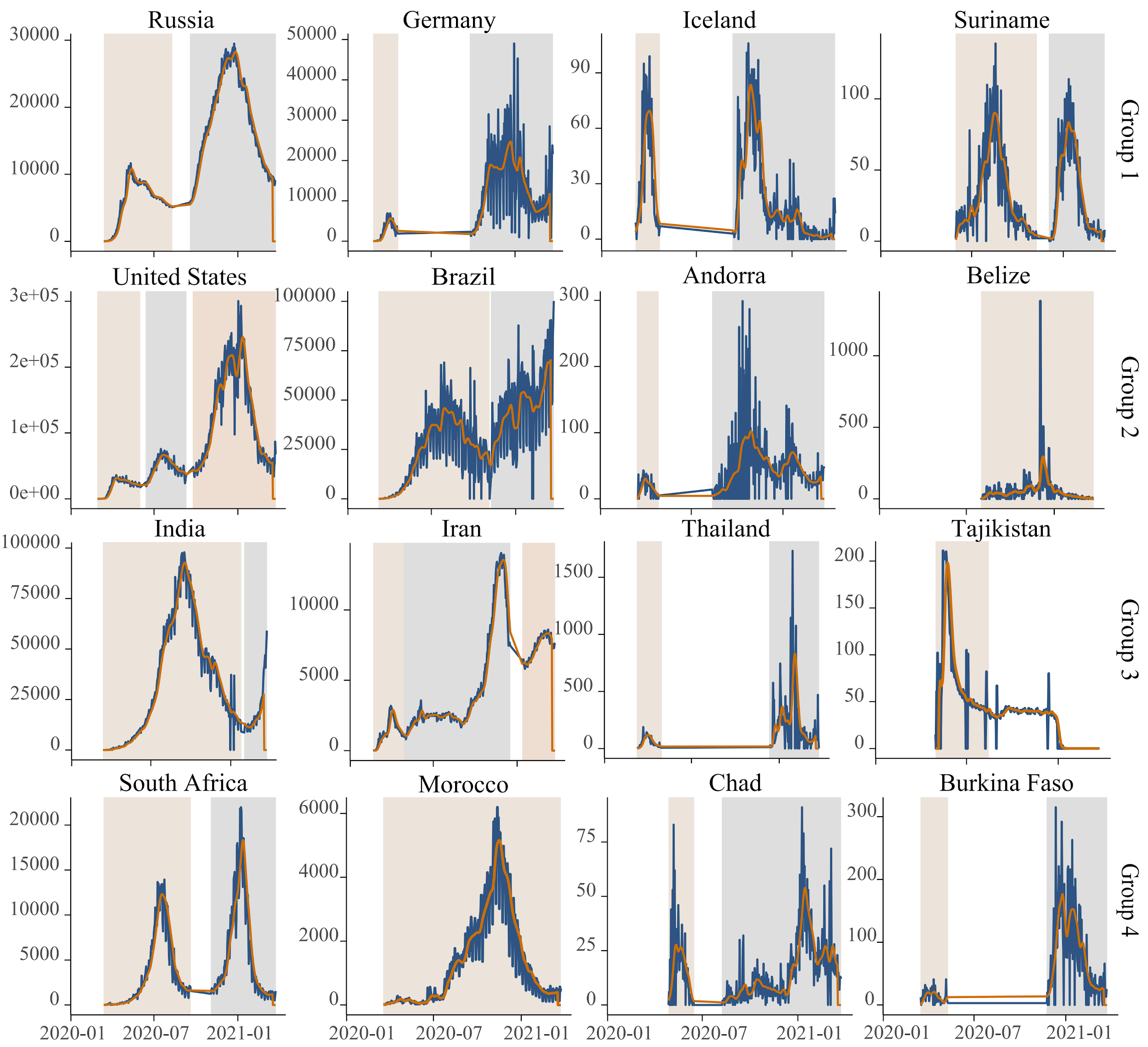

**Fig. A3.** Illustrations of raw data (in blue) and corresponding smoothed data (in orange) of daily confirmed active cases. Here we only show the top two countries and the last two countries in each country group vis-a-vis cumulative cases. The waves are depicted by the shadows with different colours.

To examine the effects of NPIs on mitigating COVID-19, we used the timing and intensity of NPIs implementations together with control variables to explain the pandemic trajectories across countries. However, the onset date of the COVID-19 outbreak varied in different countries. The dataset used in the estimation of NPIs efficacy was generated by pooling up the start date of the first, second and third wave, respectively, instead of aggregating all the countries in terms of dates. Then, we aggregated the daily time series of confirmed cases into a weekly dataset by summing all new cases in the corresponding week. To define the effect of NPIs before vaccine rollouts in our NPI effectiveness estimation, we only used case data before the date when countries started their COVID-19 vaccination programme among populations. Sensitivity analyses were also conducted to evaluate the impact of different infection-to-report delays in our estimates.

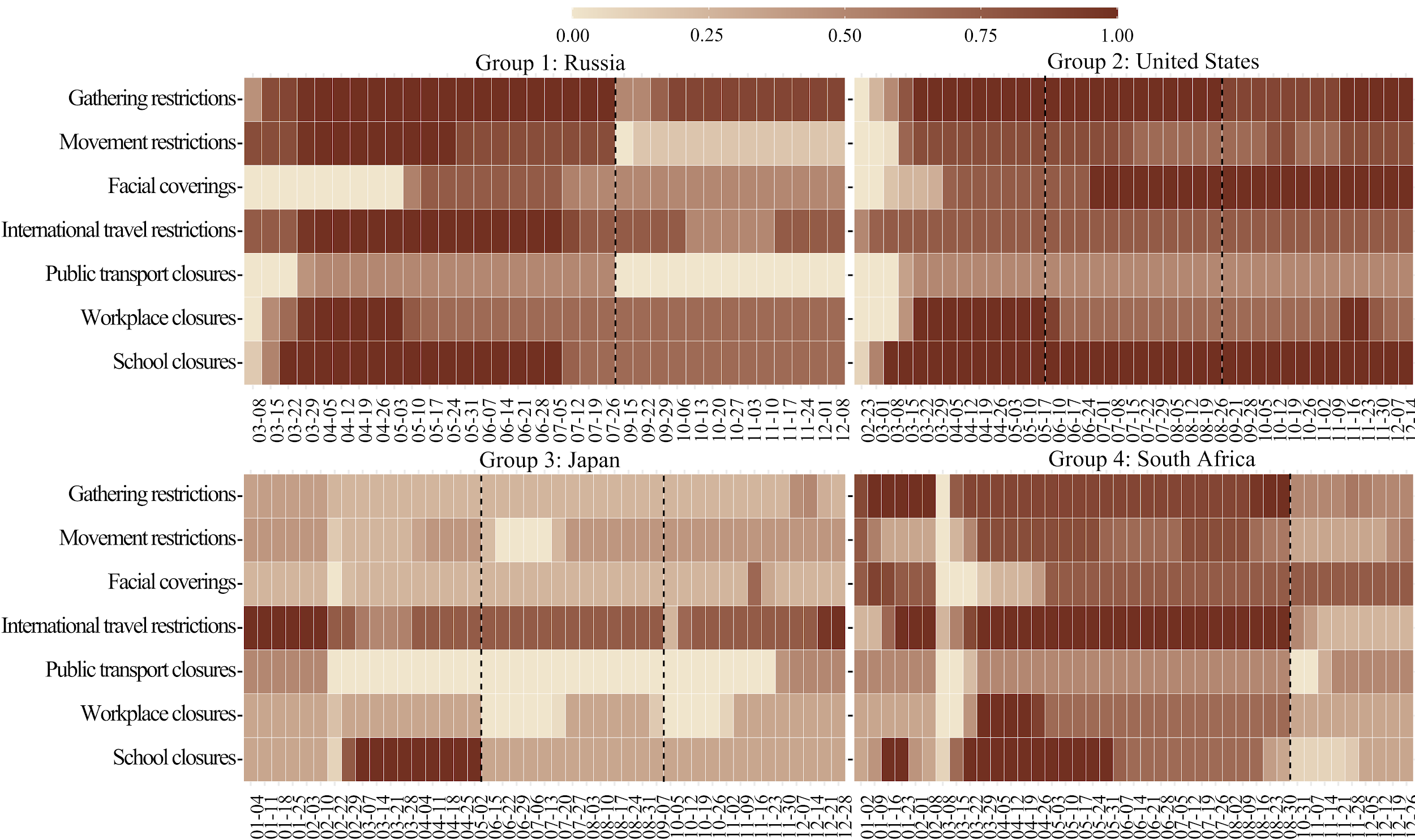

**Fig. A4.** Illustrations of timing and intensity of NPIs implementation for Russia, United States, Japan and South Africa.

*Policy data*

We employed nine NPIs (the definitions were given in Table A1) in our study, collated by the Oxford COVID-19 Government Response Tracker (OxCGRT)[14]. For each record of policy data, intensity of the nine considered NPI policies was scaled into discrete values between 0 to 1 by dividing the corresponding highest intensity value listed in Table A1, where 0 represented an absence of the NPI and 1 represented the corresponding maximum intensity. We used the timing of public and school holidays[15] to adjust the intensity of school closures to 1 on those days. Then, we integrated the NPIs policy data into the prepared datasets of weekly infection data, and the weekly intensity of each NPI was the mean of corresponding daily intensity values in each week. Finally, public events cancellations and gathering restrictions were further aggregated into a single intervention (gathering restrictions) for global and regional analysis with their average intensity for each record, and stay-at-home orders and internal movement restrictions were also aggregated into movement restrictions in the same way (for example see Fig. A4). In contrast, movement restrictions was disaggregated for national and subnational studies, while international travel restrictions was excluded due to its constant intensity across states of the US over time. The NPIs studied on each scale were given in Table A2.

**Table A1**. Definition of the employed nine NPIs from the Oxford COVID-19 Government Response Tracker (OXCGRT) in terms of their intensity.

| **NPIs** | **Intensity** | **Description** |
| --- | --- | --- |
| School closures | 1 | Closing or all schools open with alterations is recommended |
|  | 2 | Closing some levels or categories is required |
|  | 3 | Closing all levels is required |
| Workplace closures | 1 | Closing workplace is required |
|  | 2 | Closing some sectors or categories of workers is required |
|  | 3 | Closing non-essential workplaces is required |
| Public events cancellations | 1 | Cancelling public events is recommended |
|  | 2 | Cancelling public events is required |
| Gathering restrictions | 1 | Above 1000 people |
|  | 2 | Between 101-1000 people |
|  | 3 | Between 11-100 people |
|  | 4 | 10 people or less |
| Public transport closures | 1 | Closing is recommended |
|  | 2 | Closing is required |
| Stay-at-home orders | 1 | Stay at home is recommended |
|  | 2 | Stay at home with exceptions for daily exercise, grocery shopping, and 'essential' trips is required |
|  | 3 | Stay at home with minimal exceptions (e.g., allowed to leave once a week, or only one person can leave at a time, etc) is required |
| Internal movement restrictions | 1 | Travel between regions/cities is not recommended |
|  | 2 | Internal movement is restricted in place |
| International travel restrictions | 1 | Screening arrivals |
|  | 2 | Quarantine arrivals from some or all regions |
|  | 3 | Ban arrivals from some regions |
|  | 4 | Ban on all regions or total border closure |
| Facial coverings | 1 | Recommended |
|  | 2 | Required in some specified shared/public spaces outside the home with other people present, or some situations when social distancing not possible |
|  | 3 | Required in all shared/public spaces outside the home with other people present or all situations when social distancing not possible |
|  | 4 | Required outside the home at all times regardless of location or presence of other people |

Note: International travel controls were deployed for foreign travellers only, not citizens.

**Table A2**. NPIs studied on global, regional, national and subnational scales.

|  | **Analysis scale** | | | |
| --- | --- | --- | --- | --- |
| **NPIs** | **Global** | **Regional** | **National** | **Subnational** |
| School closures | 🔺 | 🔺 | 🔺 | 🔺 |
| Workplace closures | 🔺 | 🔺 | 🔺 | 🔺 |
| Public events cancellations |  |  |  |  |
| Gathering restrictions | 🔺 | 🔺 | 🔺 | 🔺 |
| Public transport closures | 🔺 | 🔺 | 🔺 | 🔺 |
| Stay-at-home orders | 🔺 | 🔺 | 🔺 | 🔺 |
| Internal movement restrictions |  |  | 🔺 | 🔺 |
| International travel restrictions | 🔺 | 🔺 |  |  |
| Facial coverings | 🔺 | 🔺 | 🔺 | 🔺 |

*Environmental and demographic covariates*

Population density is intrinsically a very factor influencing infection rate, especially for megacities[16, 17], and the elderly are more vulnerable to the pandemic due to the severe infections[18]. Different aging ratios across countries might also affect mobility and mortality[19]. The different growth rates of COVID-19 infections would also fluctuate due to disparate testing and case detection capacities[20]. The per capita health capacity may help us control this kind of variation. In addition, COVID-19 seems to possess higher transmission rates in wintertime[21].

To control for country-specific confounders in the estimates of intervention effectiveness across countries, we assembled population density, aging ratio, health capacity index, air temperature, and humidity for all these 133 study countries. We integrated these data into the datasets of confirmed cases and vaccination, respectively, in terms of corresponding documented dates. With respect to the weekly datasets, environmental conditions were aggregated into weekly with the mean value. Moreover, each environmental and demographic covariate was normalized in each dataset by dividing the corresponding maximum value independently to eliminate dimensional effects.

### B Validation and sensitivity analysis

#### **B.1** **Cross validation**

We used Cross-Validation (CV) to validate our model. We used the data of 70% countries (93), randomly sampled from 133 study countries, to build the model and estimate the overall NPI effects. To examine the performance of models, the instantaneous growth rates derived from the data of remaining 30% countries (40) were compared with the growth rates predicted by models using the corresponding implemented NPIs and country-specific characteristics in these countries. The difference between the predicted and empirically calculated instantaneous growth rates was evaluated by the mean square error (MSE). In general, MSE ranged from 0 to infinite with 0 representing the perfect prediction ability. For each of 40 countries in the validation dataset, MSE was the mean of the squared error of the instantaneous growth rates at each point of time. We used the average MSE of these 40 countries to represent the reliability of our model. This process has been repeated 50 times independently, generating 72679 pairs of predicted and empirical instantaneous growth rates, and the average MSE was (median 1.44, interquartile range [IQR] 1.25 – 2.04). Moreover, the predicted and empirical instantaneous growth rates were standardised within each country independently and respectively, by subtracting the mean and then dividing the standard error, for comparing instantaneous growth rates across countries. All the independently standardised values were shown in Fig. B1 and analysed by one-way ANOVA. The results showed that our model explained 42% variance ($var(predictions)/(var\left( predictions \right)+var\left( residuals \right))$) in the empirical instantaneous growth rates, with P value < 0.001. Fig. B2 illustrated four countries that were confronting different numbers of waves.

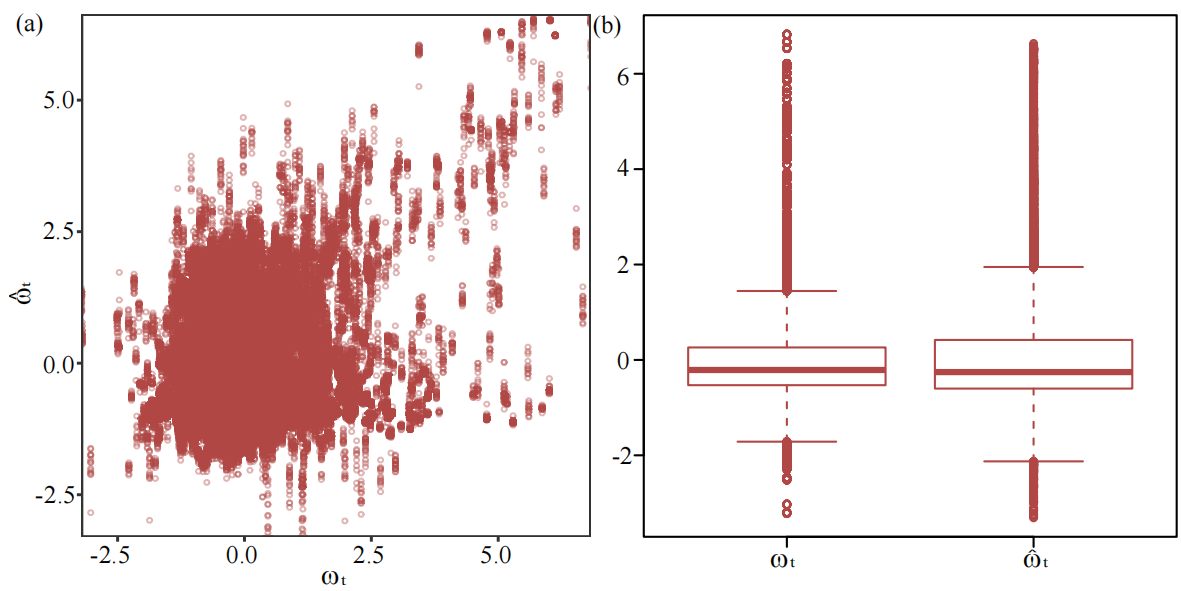

**Fig. B1.** **The results of 50 times cross-validation.** (a) The scatter plot of the standardised 76862 pairs instantaneous growth rates. (b) The box plots of the predicted ($\hat{\omega}_{t}$) and empirical ($\omega_{t}$) instantaneous growth rates, respectively. (***: p < 0.001)

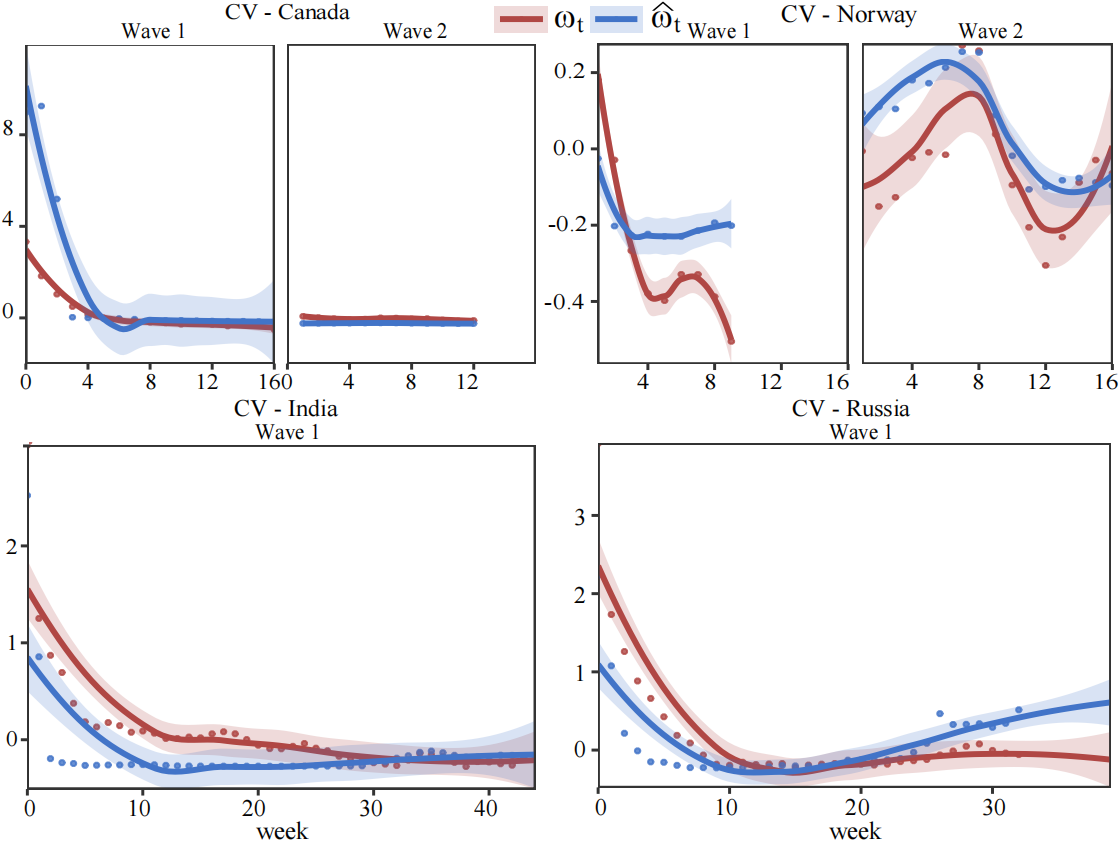

**Fig. B2.** **Comparison between the predicted instantaneous growth rates on each point of time and the empirical instantaneous growth rate.** The predicted instantaneous growth rates ($\hat{\omega}_{t}$) were represented by blue dots, where the empirical instantaneous growth rates ($\omega_{t}$) were represented by red dots. The corresponding lines were fitted by locally weighted scatterplot smoothing, with the shadowed area representing 95% confidence interval.

#### **B.2 Sensitivity analysis**

We designed three scenarios (Table B1) to perform sensitivity analyses based on our model assumptions, including 1) the probability that NPIs or vaccination performed negatively in reducing COVID-19 transmission; 2) the probability that country-specific characteristics had negative impacts on reducing COVID-19 transmission. Then, we calibrated our model with different scenarios. Results showed that the relative importance and ranks of NPIs for the reduction in $\omega_{0}$ were not significantly changed under the three scenarios (Fig. B3). For a specific policy, the decay ratio in the COVID-19 infection rate was slightly varied among three scenarios.

**Table B1.** Three scenarios in the sensitivity analysis of the model

|  | Probability in NPIs negative effect | Probability in country-specific characteristics negative effect |
| --- | --- | --- |
|  | $\alpha$ | $\beta$ |
| Scenario 1  (Lower bound) | 10% | 40% |
| Scenario 2  (Default setting) | 20% | 50% |
| Scenario 2  (Upper bound) | 30% | 60% |

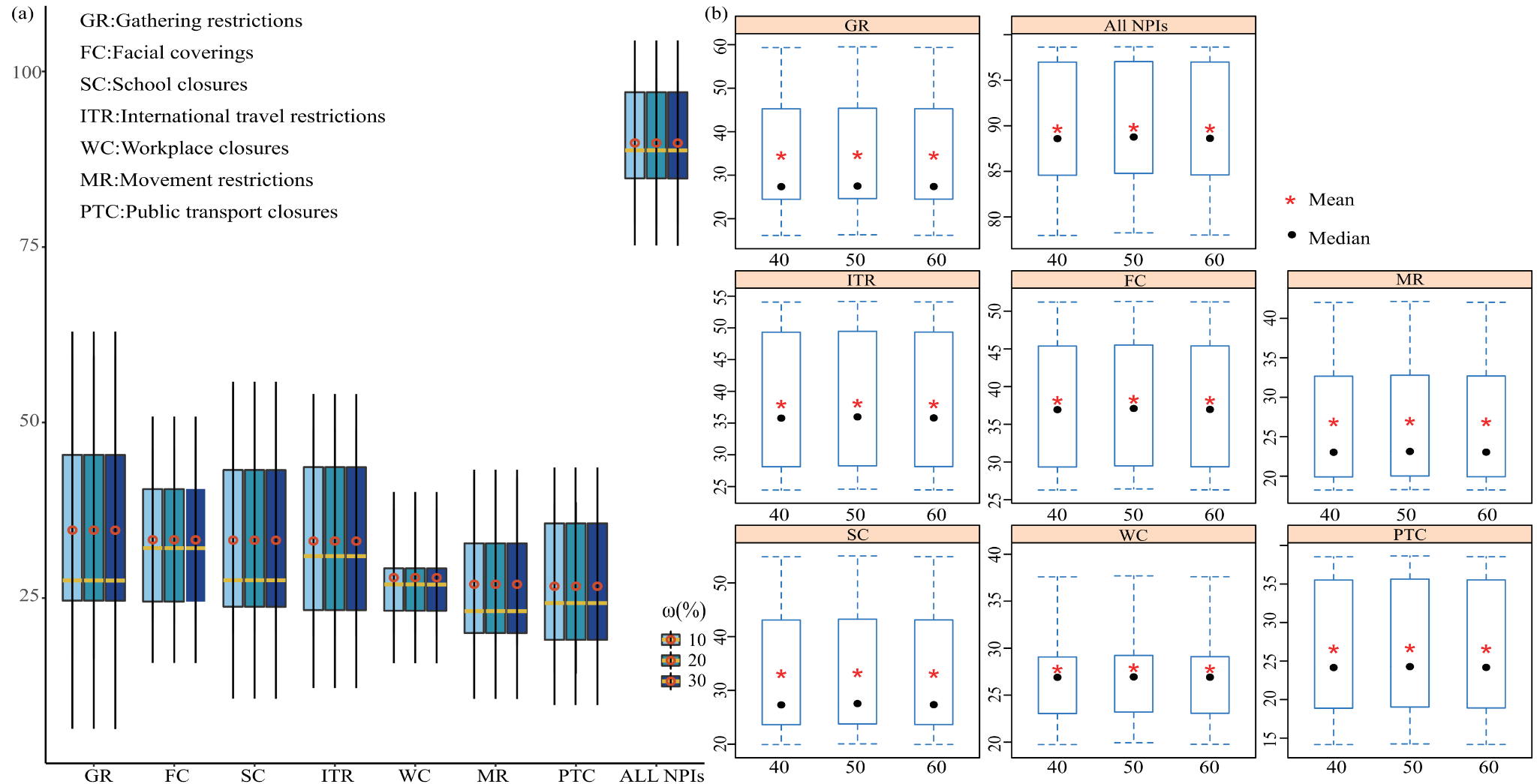

**Fig. B3.** Sensitivity analysis of the model assumption. (a) Comparison of for seven NPIs under three scenarios listed in Table B1. NPIs are ranked by the decay ratio (%$\Delta\omega_{t}$). (b) Comparison of the three scenarios for the efficacy of individual NPIs.

#### **B.3 MCMC convergence**

We calibrated our model with the Markov chain Monte Carlo (MCMC) sampling algorithm. We used R-hat statistics and relative effective sample size to present MCMC performance during our model calibration. The results showed that our model calibrations with MCMC had a good convergence (see Fig. B6(a)) and the sample size was effective in decomposing the variation in decay ratios into NPI effectiveness (see Fig. B6(b)).

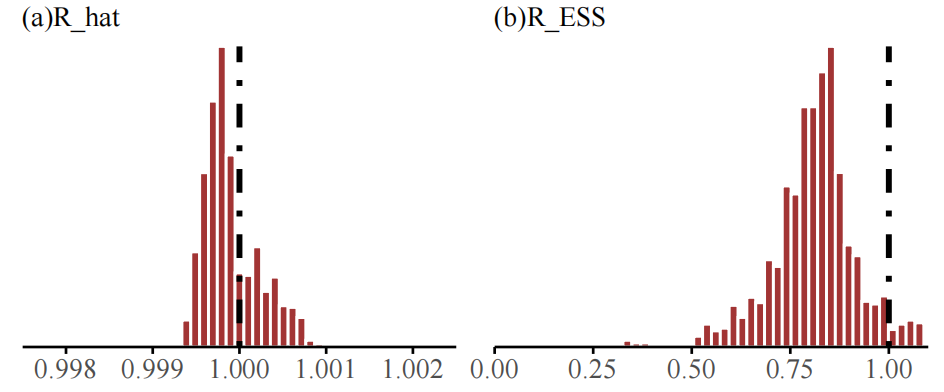

**Fig. B4.** (a)R-hat statistic taken from a run using the default model with default settings and values for all parameters. Values are close to 1, indicating convergence. (b) Relative effective sample size taken from a run using the default model with default settings and values for all parameters. Value 1 indicates perfect decorrelation between samples. Values above (below) 1 indicate that the effective number of samples is higher (lower) than the actual number of samples due to negative (positive) correlation, respectively.

### C Additional results

#### **C.1 Collinearity**

Nine common NPIs were analysed in this study, including school closures, workplace closures, public events cancellations, gathering restrictions, public transport closures, stay-at-home orders, internal movement restrictions, international travel restrictions, and facial coverings. To avoid the multicollinearity caused by high correlations and constraints in models to separately identify the NPI effect, we conducted a correlation analysis of NPI variables above. As shown in Fig. C1, NPIs having some causal meanings did have higher correlation coefficients (about 0.5). For this reason, we defined a “movement restrictions” variable using the average of intensities of stay-at-home orders and internal movement restrictions, and generated a new “gathering restrictions” variable from the average of intensities of public events cancellations and gathering restrictions.

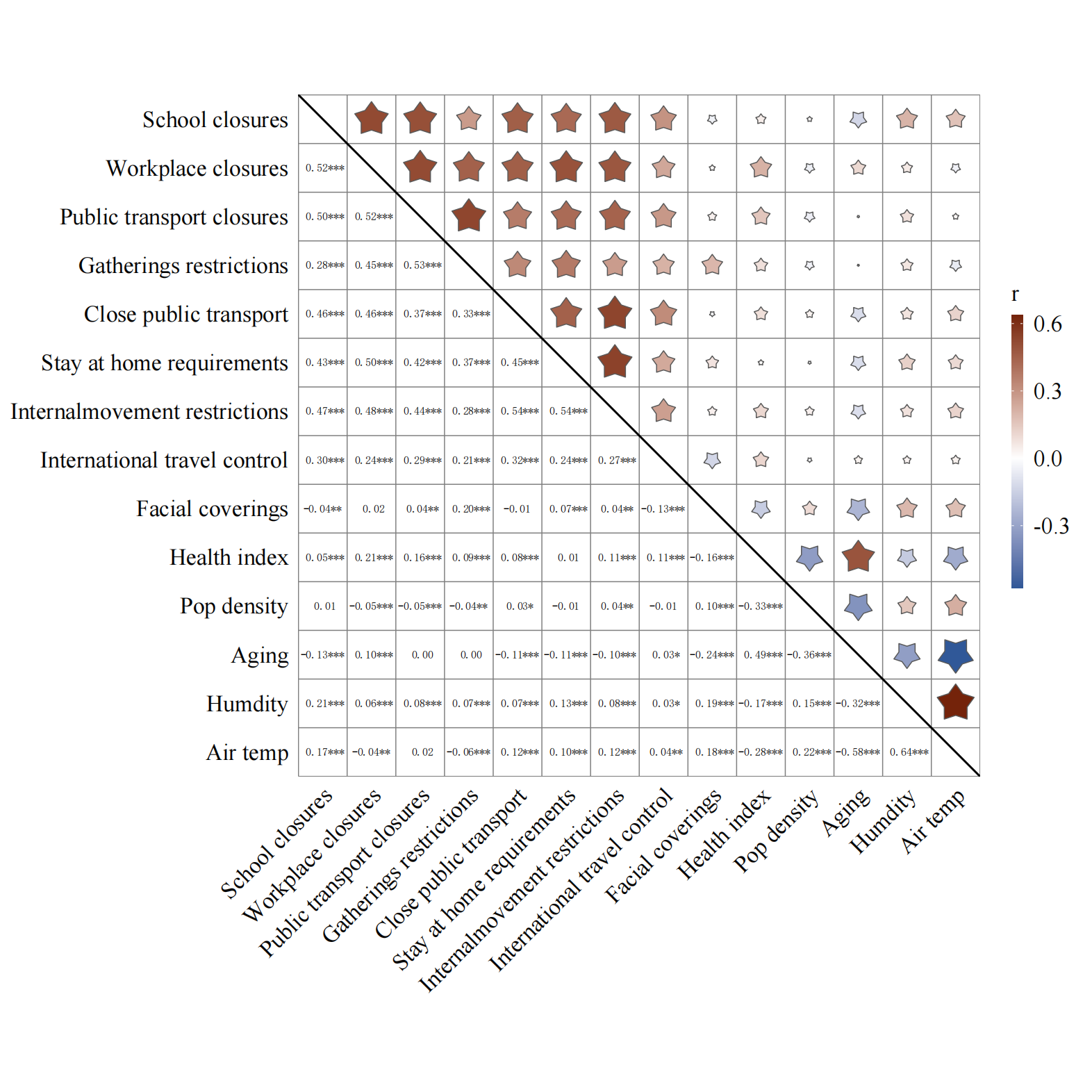

**Fig. C1.** Correlations between all collected explanatory variables in initial. (*: p < 0.05; **: p < 0.01; ***: p < 0.001).

#### **C.2 Country groups and waves**

To differentiate the performance of NPIs against COVID-19 across countries and waves, we divided the 133 study countries into four country groups based on their morbidity and mortality of COVID-19 together with geographical proximity (see Fig. C2) and then defined waves for each country (Table C1 – C4). In fact, the capacity of testing and diagnosis was a major confounder in different NPI effects across countries. However, the testing rate was not generally available for all the 133 countries. We used the number of testing per thousand people on 22 June 2021 to represent the testing rate in each country having testing data, where the morbidity and mortality of this country were represented by the total cases/deaths per million people at that day, respectively. Then, we tested Pearson's product-moment correlation between the total tests per thousand people and the total cases/deaths per million people, respectively (see Table C5). The results showed that the testing rate was correlated to the morbidity and mortality indicating we divided 133 countries into four country groups implicitly by the testing rate.

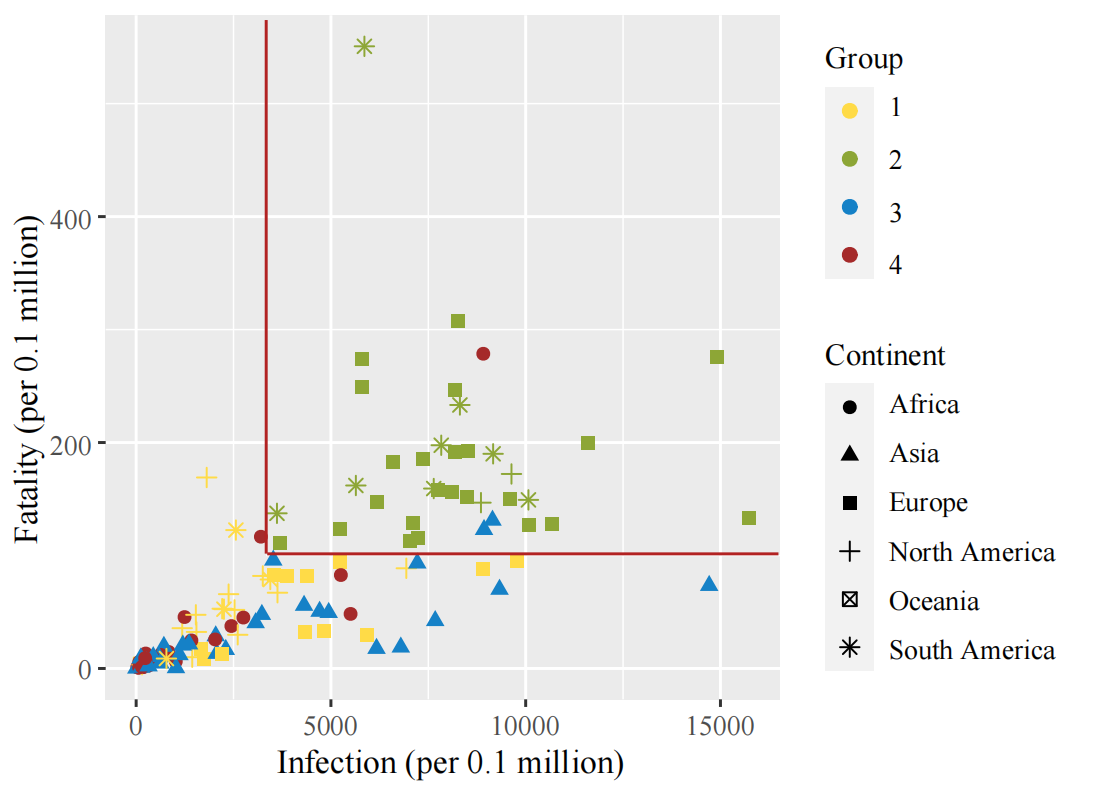

**Fig. C2.** Distribution of the 133 study countries in terms of morbidity and mortality. The red lines represent the grading thresholds for high morbidity and mortality.

**Table C1**. The start date and end date of each wave in each country of Group 1.

| **Country** | **Start Date (dd/mm/yy)** | | | **End Date (dd/mm/yy)** | | |
| --- | --- | --- | --- | --- | --- | --- |
|  | **Wave 1** | **Wave 2** | **Wave 3** | **Wave 1** | **Wave 2** | **Wave 3** |
| Albania | 04/24/20 | 01/09/21 |  | 01/04/21 | 03/25/21 |  |
| Australia | 03/14/20 | 06/20/20 |  | 04/19/20 | 09/18/20 |  |
| Austria | 03/10/20 | 09/01/20 |  | 05/01/20 | 03/25/21 |  |
| Bahamas | 07/22/20 |  |  | 02/01/21 |  |  |
| Belarus | 04/02/20 | 08/16/20 |  | 08/11/20 | 03/25/21 |  |
| Belize | 08/08/20 |  |  | 03/25/21 |  |  |
| Bolivia | 04/13/20 | 11/30/20 |  | 11/01/20 | 03/25/21 |  |
| Canada | 03/14/20 | 09/20/20 |  | 06/20/20 | 03/25/21 |  |
| Costa Rica | 03/26/20 |  |  | 03/25/21 |  |  |
| Croatia | 03/19/20 | 10/10/20 |  | 04/24/20 | 03/25/21 |  |
| Cuba | 03/28/20 | 07/31/20 |  | 05/20/20 | 03/25/21 |  |
| Cyprus | 03/24/20 | 10/13/20 | 02/13/21 | 04/25/20 | 02/04/21 | 03/25/21 |
| Denmark | 03/10/20 | 07/27/20 |  | 05/10/20 | 03/25/21 |  |
| Dominican Republic | 03/23/20 | 11/01/20 |  | 09/05/20 | 03/25/21 |  |
| Ecuador | 03/19/20 |  |  | 03/25/21 |  |  |
| El Salvador | 05/01/20 | 09/20/20 |  | 09/15/20 | 03/25/21 |  |
| Estonia | 03/14/20 | 10/13/20 |  | 04/30/20 | 03/25/21 |  |
| Finland | 03/13/20 | 08/05/20 |  | 06/07/20 | 03/25/21 |  |
| Germany | 03/03/20 | 10/01/20 |  | 04/20/20 | 03/25/21 |  |
| Greece | 03/11/20 | 08/10/20 | 01/22/21 | 04/10/20 | 01/11/21 | 03/25/21 |
| Guatemala | 04/22/20 |  |  | 03/25/21 |  |  |
| Guyana | 08/02/20 |  |  | 03/25/21 |  |  |
| Haiti | 05/12/20 | 12/03/20 |  | 10/05/20 | 03/25/21 |  |
| Honduras | 04/03/20 | 02/04/21 |  | 01/12/21 | 03/25/21 |  |
| Iceland | 03/13/20 | 09/16/20 |  | 04/22/20 | 12/23/20 |  |
| Ireland | 03/17/20 | 09/01/20 | 12/10/20 | 05/23/20 | 11/10/20 | 03/25/21 |
| Jamaica | 04/18/20 | 08/15/20 | 12/28/20 | 05/10/20 | 10/30/20 | 03/25/21 |
| Latvia | 03/20/20 | 09/25/20 |  | 04/15/20 | 03/25/21 |  |
| Lithuania | 03/22/20 | 10/01/20 |  | 05/01/20 | 03/25/21 |  |
| Mexico | 03/20/20 | 11/15/20 |  | 11/01/20 | 03/25/21 |  |
| Norway | 03/06/20 | 09/01/20 | 02/15/21 | 05/10/20 | 01/18/21 | 03/25/21 |
| Paraguay | 05/04/20 | 11/07/20 |  | 10/20/20 | 03/25/21 |  |
| Poland | 03/17/20 | 02/03/21 |  | 01/12/21 | 03/25/21 |  |
| Portugal | 03/15/20 | 09/20/20 | 12/23/20 | 05/10/20 | 12/13/21 | 03/25/21 |
| Russia | 03/20/20 | 09/25/20 |  | 08/10/20 | 03/25/21 |  |
| Suriname | 06/06/20 | 12/10/20 |  | 11/09/20 | 03/25/21 |  |
| Trinidad and Tobago | 08/09/20 |  |  | 03/25/21 |  |  |
| Ukraine | 03/26/20 | 02/13/21 |  | 02/05/21 | 03/25/21 |  |
| Uruguay | 03/23/20 |  |  | 03/25/21 |  |  |
| Venezuela | 05/18/20 | 01/02/21 |  | 12/20/20 | 03/25/21 |  |

**Table C2**. The start date and end date of each wave in each country of Group 2.

| **Country** | **Start Date (dd/mm/yy)** | | | **End Date (dd/mm/yy)** | | |
| --- | --- | --- | --- | --- | --- | --- |
|  | **Wave 1** | **Wave 2** | **Wave 3** | **Wave 1** | **Wave 2** | **Wave 3** |
| Andorra | 03/21/20 | 08/19/20 |  | 04/27/20 | 03/25/21 |  |
| Argentina | 03/22/20 |  |  | 03/25/21 |  |  |
| Belgium | 03/08/20 | 08/10/20 |  | 05/10/20 | 01/03/21 |  |
| Bosnia and Herzegovina | 03/27/20 | 10/10/20 |  | 10/01/20 | 03/25/21 |  |
| Brazil | 03/15/20 | 11/15/20 |  | 11/05/20 | 03/25/21 |  |
| Bulgaria | 03/18/20 | 10/10/20 | 02/01/21 | 08/25/20 | 01/15/21 | 03/25/21 |
| Chile | 03/17/20 |  |  | 03/25/21 |  |  |
| Colombia | 03/21/20 | 10/01/20 |  | 10/01/20 | 03/25/21 |  |
| Czechia | 03/12/20 | 09/20/20 | 12/04/20 | 04/20/20 | 11/23/20 | 03/25/21 |
| France | 03/03/20 | 08/10/20 |  | 04/25/20 | 03/25/21 |  |
| Hungary | 03/21/20 | 09/06/20 |  | 05/15/20 | 03/25/21 |  |
| Italy | 02/24/20 | 10/01/20 | 02/10/21 | 06/10/20 | 01/25/21 | 03/25/21 |
| Luxembourg | 03/16/20 | 10/05/20 |  | 04/21/20 | 03/25/21 |  |
| Moldova | 03/28/20 | 01/17/21 |  | 12/28/20 | 03/25/21 |  |
| Netherlands | 03/07/20 | 07/30/20 | 11/29/20 | 05/20/20 | 11/03/20 | 03/25/21 |
| Panama | 03/21/20 | 11/15/20 |  | 11/01/20 | 03/25/21 |  |
| Peru | 03/19/20 | 07/15/20 | 01/02/21 | 07/01/20 | 12/04/20 | 03/25/21 |
| Slovenia | 03/12/20 | 09/25/20 |  | 04/15/20 | 03/25/21 |  |
| Spain | 03/04/20 | 07/13/20 |  | 04/30/20 | 03/25/21 |  |
| Sweden | 03/07/20 | 09/25/20 |  | 07/05/20 | 03/25/21 |  |
| Switzerland | 03/06/20 | 10/05/20 |  | 04/23/20 | 03/25/21 |  |
| United Kingdom | 03/04/20 | 09/10/20 | 12/01/20 | 06/10/20 | 11/22/20 | 03/25/21 |
| United States | 03/06/20 | 06/20/20 | 10/01/20 | 06/01/20 | 09/10/20 | 03/25/21 |

**Table C3**. The start date and end date of each wave in each country of Group 3.

| **Country** | **Start Date (dd/mm/yy)** | | | **End Date (dd/mm/yy)** | | |
| --- | --- | --- | --- | --- | --- | --- |
|  | **Wave 1** | **Wave 2** | **Wave 3** | **Wave 1** | **Wave 2** | **Wave 3** |
| Afghanistan | 04/03/20 | 11/20/20 |  | 08/15/20 | 03/25/21 |  |
| Azerbaijan | 03/30/20 | 10/10/20 |  | 08/05/20 | 03/25/21 |  |
| Bahrain | 03/24/20 | 08/31/20 | 11/27/20 | 08/10/20 | 11/10/20 | 03/25/21 |
| Bangladesh | 04/08/20 | 11/15/20 |  | 10/01/20 | 03/25/21 |  |
| China | 01/29/20 | 11/13/20 |  | 03/10/20 | 02/01/21 |  |
| Georgia | 09/05/20 |  |  | 03/25/21 |  |  |
| India | 03/20/20 | 02/08/21 |  | 01/25/21 | 03/25/21 |  |
| Indonesia | 03/18/20 |  |  | 03/25/21 |  |  |
| Iran | 02/27/20 | 05/05/20 | 01/19/21 | 04/30/20 | 12/17/20 | 03/25/21 |
| Iraq | 03/24/20 | 01/15/21 |  | 12/26/20 | 03/25/21 |  |
| Israel | 03/16/20 | 06/01/20 | 11/27/20 | 05/08/20 | 11/01/20 | 03/25/21 |
| Japan | 03/05/20 | 06/25/20 | 10/15/20 | 05/17/20 | 09/25/20 | 03/25/21 |
| Jordan | 03/28/20 | 01/22/21 |  | 01/09/21 | 03/25/21 |  |
| Kazakhstan | 03/29/20 |  |  | 09/10/20 |  |  |
| Kuwait | 04/03/20 | 12/26/20 |  | 11/22/20 | 03/25/21 |  |
| Kyrgyzstan | 04/09/20 | 10/01/20 |  | 08/26/20 | 03/25/21 |  |
| Lebanon | 03/24/20 |  |  | 03/25/21 |  |  |
| Malaysia | 03/09/20 | 10/01/20 |  | 06/13/20 | 03/25/21 |  |
| Myanmar | 08/26/20 |  |  | 01/29/21 |  |  |
| Nepal | 05/15/20 | 07/30/20 |  | 07/10/20 | 03/25/21 |  |
| Oman | 04/06/20 | 08/27/20 |  | 08/09/20 | 03/25/21 |  |
| Pakistan | 03/16/20 | 10/26/20 |  | 09/01/20 | 03/25/21 |  |
| Philippines | 03/18/20 | 01/01/21 |  | 11/14/20 | 03/25/21 |  |
| Qatar | 03/12/20 | 01/10/21 |  | 08/10/20 | 03/25/21 |  |
| Saudi Arabia | 03/18/20 |  |  | 03/25/21 |  |  |
| Singapore | 03/20/20 | 07/13/20 |  | 06/28/20 | 09/15/20 |  |
| South Korea | 02/21/20 | 08/10/20 | 11/10/20 | 04/20/20 | 09/30/20 | 03/25/21 |
| Sri Lanka | 04/21/20 | 10/03/20 |  | 07/21/20 | 03/25/21 |  |
| Syria | 07/04/20 | 10/01/20 |  | 09/20/20 | 03/25/21 |  |
| Tajikistan | 05/07/20 |  |  | 08/15/20 |  |  |
| Thailand | 03/16/20 | 12/18/20 |  | 04/30/20 | 03/25/21 |  |
| Turkey | 03/20/20 | 11/21/20 | 01/23/21 | 05/25/20 | 01/12/21 | 03/25/21 |
| United Arab Emirates | 03/25/20 | 08/15/20 |  | 08/08/20 | 03/25/21 |  |
| Uzbekistan | 04/03/20 |  |  | 12/01/20 |  |  |

**Table C4**. The start date and end date of each wave in each country of Group 4.

| **Country** | **Start Date (dd/mm/yy)** | | | **End Date (dd/mm/yy)** | | |
| --- | --- | --- | --- | --- | --- | --- |
|  | **Wave 1** | **Wave 2** | **Wave 3** | **Wave 1** | **Wave 2** | **Wave 3** |
| Algeria | 03/23/20 | 10/14/20 |  | 10/13/20 | 03/25/21 |  |
| Angola | 06/29/20 |  |  | 03/25/21 |  |  |
| Benin | 05/07/20 | 06/08/20 |  | 05/21/20 | 03/25/21 |  |
| Botswana | 06/30/20 | 10/20/20 |  | 10/05/20 | 03/25/21 |  |
| Burkina Faso | 03/23/20 | 12/01/20 |  | 05/10/20 | 03/25/21 |  |
| Cameroon | 04/01/20 |  |  | 03/05/21 |  |  |
| Cape Verde | 05/06/20 | 12/28/20 |  | 12/01/20 | 03/25/21 |  |
| Central African Republic | 05/14/20 |  |  | 08/05/20 |  |  |
| Chad | 05/04/20 | 08/15/20 |  | 06/15/20 | 03/25/21 |  |
| Congo | 04/18/20 | 12/01/20 |  | 09/20/20 | 03/25/21 |  |
| Cote d'Ivoire | 04/07/20 | 12/19/20 |  | 09/05/20 | 03/25/21 |  |
| Djibouti | 04/08/20 | 05/09/20 |  | 05/06/20 | 07/01/20 |  |
| Egypt | 03/19/20 | 10/20/20 |  | 08/20/20 | 03/25/21 |  |
| Eswatini | 06/10/20 | 12/01/20 |  | 10/30/20 | 03/25/21 |  |
| Ethiopia | 05/22/20 | 01/15/21 |  | 12/29/20 | 03/25/21 |  |
| Gabon | 04/21/20 | 01/01/21 |  | 08/15/20 | 03/25/21 |  |
| Gambia | 07/23/20 | 01/23/21 |  | 09/30/20 | 03/25/21 |  |
| Ghana | 04/10/20 | 10/17/20 |  | 09/20/20 | 03/25/21 |  |
| Guinea | 04/05/20 |  |  | 03/25/21 |  |  |
| Kenya | 05/06/20 | 10/07/20 | 02/15/21 | 09/01/20 | 01/06/21 | 03/25/21 |
| Libya | 06/09/20 |  |  | 03/25/21 |  |  |
| Madagascar | 05/20/20 |  |  | 10/18/20 |  |  |
| Malawi | 05/28/20 | 12/15/20 |  | 09/05/20 | 03/25/21 |  |
| Mauritania | 05/19/20 | 11/01/20 |  | 10/10/20 | 03/25/21 |  |
| Morocco | 03/25/20 |  |  | 03/25/21 |  |  |
| Mozambique | 06/01/20 | 12/20/20 |  | 11/04/20 | 03/25/21 |  |
| Namibia | 06/27/20 | 11/13/20 |  | 10/20/20 | 03/25/21 |  |
| Nigeria | 04/16/20 | 11/30/20 |  | 09/20/20 | 03/25/21 |  |
| Senegal | 03/27/20 | 11/25/20 |  | 11/01/20 | 03/25/21 |  |
| Somalia | 04/17/20 | 08/20/20 | 02/01/21 | 08/05/20 | 12/25/20 | 03/25/21 |
| South Africa | 03/20/20 | 11/10/20 |  | 09/20/20 | 03/25/21 |  |
| Sudan | 04/21/20 | 11/05/20 |  | 09/15/20 | 03/25/21 |  |
| Tunisia | 03/27/20 |  |  | 03/25/21 |  |  |
| Uganda | 05/30/20 |  |  | 03/25/21 |  |  |
| Zambia | 05/08/20 | 12/01/20 |  | 11/02/20 | 03/25/21 |  |
| Zimbabwe | 05/29/20 | 11/10/20 |  | 09/20/20 | 03/25/21 |  |

**Table C5.** Pearson's product-moment correlation between the total tests per thousand people and the total cases/deaths per million people, respectively.

|  | **Correlation** | **P value** |
| --- | --- | --- |
| The total cases per million people | 0.5307  (95% CI, 0.3821, 0.6525) | 2.089e-09 |
| The total deaths per million people | 0.2527  (95%CI, 0.0696,0.4193) | 0.007457 |

#### **C.3 The USA states groups and waves**

The USA states were divided into three groups in the similar way as shown in Section C.2. Table C6 – C8 show specific states in each group and their detailed waves periods.

**Table C6**. The start date and end date of each wave in each state of Group 1.

| **State** | **Start Date (dd/mm/yy)** | | | **End Date (dd/mm/yy)** | | |
| --- | --- | --- | --- | --- | --- | --- |
|  | **Wave 1** | **Wave 2** | **Wave 3** | **Wave 1** | **Wave 2** | **Wave 3** |
| Alaska | 03/06/20 | 06/20/20 | 09/01/20 | 06/01/20 | 08/20/20 | 06/20/21 |
| Hawaii | 03/06/20 | 05/20/20 | 10/10/20 | 05/10/20 | 10/01/20 | 06/20/21 |
| Maine | 03/06/20 | 07/15/20 | 10/01/20 | 07/10/20 | 09/10/20 | 06/20/21 |
| New Hampshire | 03/06/20 | 06/20/20 | 09/01/20 | 06/01/20 | 08/20/20 | 06/20/21 |
| New York | 03/06/20 | 06/10/20 | 08/25/20 | 06/01/20 | 08/10/20 | 06/20/21 |
| Oregon | 03/06/20 | 05/20/20 | 09/10/20 | 05/10/20 | 09/01/20 | 06/20/21 |
| Utah | 03/06/20 | 05/15/20 | 08/20/20 | 05/10/20 | 08/10/20 | 06/20/21 |
| Vermont | 03/06/20 | 05/20/20 | 10/01/20 | 05/10/20 | 09/10/20 | 06/20/21 |
| Washington | 03/06/20 | 05/20/20 | 09/05/20 | 05/01/20 | 08/25/20 | 06/20/21 |

**Table C7**. The start date and end date of each wave in each state of Group 2.

| **State** | **Start Date (dd/mm/yy)** | | | **End Date (dd/mm/yy)** | | |
| --- | --- | --- | --- | --- | --- | --- |
|  | **Wave 1** | **Wave 2** | **Wave 3** | **Wave 1** | **Wave 2** | **Wave 3** |
| California | 03/06/20 | 10/01/20 |  | 09/10/20 | 06/20/21 |  |
| Colorado | 03/06/20 | 06/05/20 | 09/01/20 | 05/30/20 | 08/22/20 | 06/20/21 |
| District of Columbia | 03/06/20 | 06/20/20 | 10/01/20 | 06/10/20 | 09/10/20 | 06/20/21 |
| Delaware | 03/06/20 | 06/10/20 | 08/01/20 | 06/01/20 | 07/20/20 | 06/20/21 |
| Florida | 03/06/20 | 05/20/20 | 10/01/20 | 05/10/20 | 09/10/20 | 06/20/21 |
| Idaho | 03/06/20 | 06/01/20 | 09/01/20 | 05/01/20 | 08/25/20 | 06/20/21 |
| Kansas | 03/06/20 | 06/10/20 | 09/15/20 | 05/30/20 | 09/01/20 | 06/20/21 |
| Kentucky | 03/06/20 | 06/15/20 | 09/15/20 | 06/01/20 | 09/01/20 | 06/20/21 |
| Maryland | 03/06/20 | 06/20/20 | 10/01/20 | 06/05/20 | 09/10/20 | 06/20/21 |
| Minnesota | 03/06/20 | 06/20/20 | 09/05/20 | 06/01/20 | 08/25/20 | 06/20/21 |
| Missouri | 03/06/20 | 06/01/20 |  | 05/15/20 | 06/20/21 |  |
| Montana | 03/06/20 | 06/01/20 | 08/20/20 | 05/10/20 | 08/05/20 | 06/20/21 |
| North Carolina | 03/06/20 | 08/01/20 | 09/10/20 | 07/25/20 | 08/22/20 | 06/20/21 |
| Nebraska | 03/06/20 | 06/20/20 | 08/15/20 | 06/10/20 | 08/10/20 | 06/20/21 |
| Nevada | 03/06/20 | 06/01/20 | 09/15/20 | 05/15/20 | 09/01/20 | 06/20/21 |
| Ohio | 03/06/20 | 06/10/20 | 10/01/20 | 06/01/20 | 09/10/20 | 06/20/21 |
| Texas | 03/06/20 | 09/05/20 | 10/05/20 | 08/25/20 | 09/25/20 | 06/20/21 |
| Virginia | 03/06/20 | 06/20/20 | 09/20/20 | 06/10/20 | 09/10/20 | 06/20/21 |
| West Virginia | 03/06/20 | 06/10/20 | 08/20/20 | 06/01/20 | 08/10/20 | 06/20/21 |
| Wisconsin | 03/06/20 | 06/20/20 | 08/15/20 | 06/10/20 | 08/05/20 | 06/20/21 |
| Wyoming | 03/06/20 | 06/10/20 | 09/01/20 | 05/22/20 | 08/20/20 | 06/20/21 |

**Table C8**. The start date and end date of each wave in each state of Group 3.

| **State** | **Start Date (dd/mm/yy)** | | | **End Date (dd/mm/yy)** | | |
| --- | --- | --- | --- | --- | --- | --- |
|  | **Wave 1** | **Wave 2** | **Wave 3** | **Wave 1** | **Wave 2** | **Wave 3** |
| Alabama | 03/06/20 | 09/20/20 |  | 09/15/20 | 06/20/21 |  |
| Arkansas | 03/06/20 | 05/01/20 | 08/15/20 | 04/25/20 | 08/05/20 | 06/20/21 |
| Arizona | 03/06/20 | 09/10/20 |  | 09/01/20 | 06/20/21 |  |
| Connecticut | 03/06/20 | 06/20/20 | 08/20/20 | 06/01/20 | 08/05/20 | 06/20/21 |
| Georgia | 03/06/20 | 05/20/20 | 10/01/20 | 05/10/20 | 09/15/20 | 06/20/21 |
| Iowa | 03/06/20 | 06/10/20 | 09/10/20 | 06/01/20 | 08/25/20 | 06/20/21 |
| Illinois | 03/06/20 | 06/20/20 | 09/20/20 | 06/01/20 | 09/01/20 | 06/20/21 |
| Indiana | 03/06/20 | 06/20/20 | 10/01/20 | 06/01/20 | 09/10/20 | 06/20/21 |
| Louisiana | 03/06/20 | 05/20/20 | 09/20/20 | 05/10/20 | 09/10/20 | 06/20/21 |
| Massachusetts | 03/06/20 | 06/20/20 |  | 06/01/20 | 06/20/21 |  |
| Michigan | 03/06/20 | 05/25/20 | 10/01/20 | 05/15/20 | 09/20/20 | 06/20/21 |
| Mississippi | 03/06/20 | 09/10/20 |  | 09/01/20 | 06/20/21 |  |
| North Dakota | 03/06/20 | 06/20/20 |  | 06/01/20 | 06/20/21 |  |
| New Jersey | 03/06/20 | 06/30/20 | 08/15/20 | 06/01/20 | 08/05/20 | 06/20/21 |
| New Mexico | 03/06/20 | 06/10/20 | 09/01/20 | 06/01/20 | 08/22/20 | 06/20/21 |
| Oklahoma | 03/06/20 | 05/10/20 | 08/01/20 | 05/02/20 | 07/25/20 | 06/20/21 |
| Pennsylvania | 03/06/20 | 06/10/20 | 09/20/20 | 06/01/20 | 09/10/20 | 06/20/21 |
| Rhode Island | 03/06/20 | 06/25/20 | 09/01/20 | 06/10/20 | 08/20/20 | 06/20/21 |
| South Carolina | 03/06/20 | 05/15/20 | 10/01/20 | 05/02/20 | 09/10/20 | 06/20/21 |
| South Dakota | 03/06/20 | 06/20/20 |  | 06/01/20 | 06/20/21 |  |
| Tennessee | 03/06/20 | 05/16/20 | 09/02/20 | 05/10/20 | 08/25/20 | 06/20/21 |

#### **C.4 Global efficacy of COVID-19 interventions differing in the three waves**

The differences of NPI effects among three waves were tested using a Wilcoxon signed-rank test, a non-parametric statistical hypothesis test for comparing NPIs effects between pairs of the three waves (see Fig. C3). The results showed that the relative effects of NPIs were significantly changed between waves.

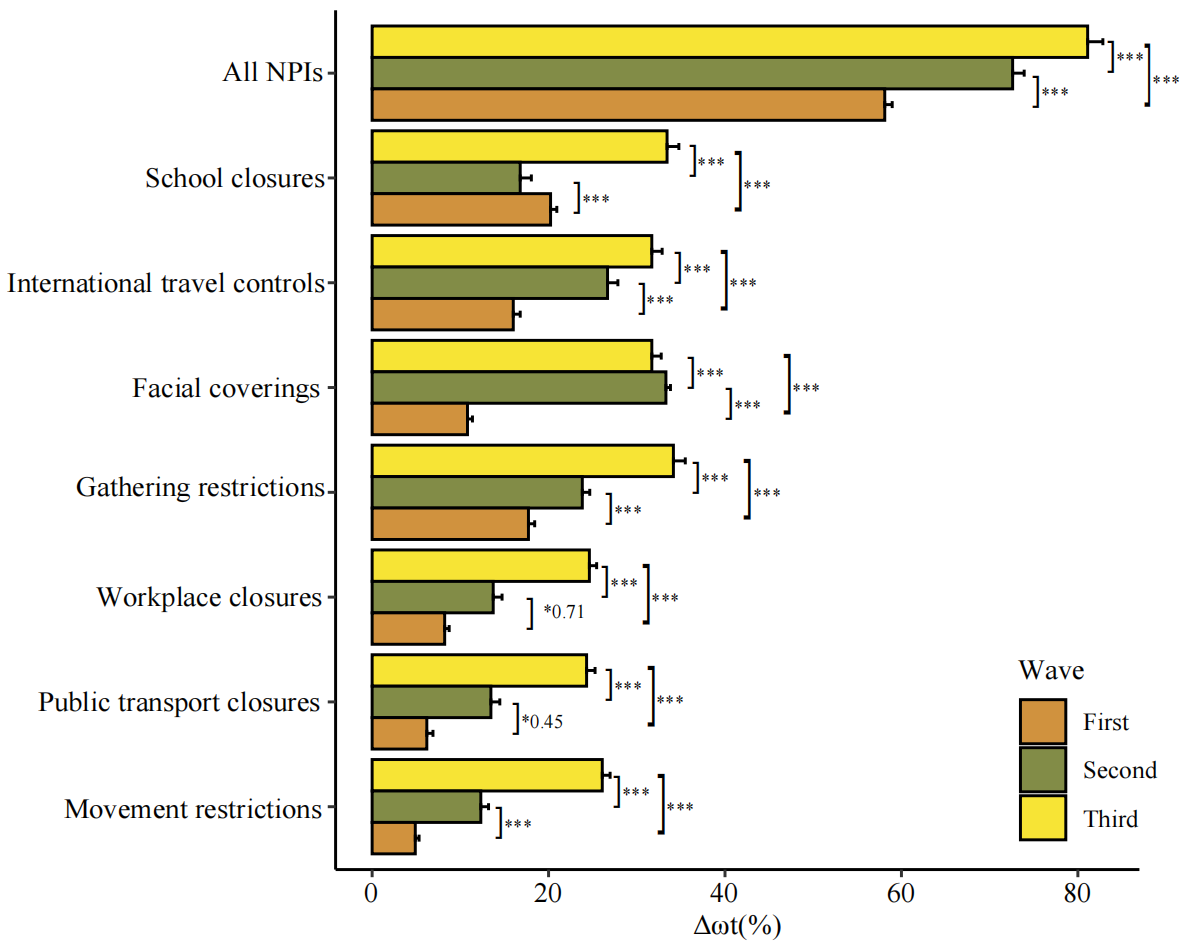

**Fig. C3.** **Difference of global NPI effects among the three COVID-19 waves.** The comparison indicates that effects of policies were significantly changed between waves. In general, effects of policies in the second wave were higher than those in the first wave, and effects in the third wave were higher than those in the second wave. In the first wave, COVID-19 interventions with relatively high effects included gathering restrictions, facial coverings, and school closures. In the second wave, the primary effective interventions were workplace closures, facial coverings, and international travel restrictions. In the third wave, the major NPIs with effectiveness were gathering restrictions, international travel restrictions, and school closures. NPIs were ranked by the decay ratio in the COVID-19 infection rate estimated in the first wave. (*: p < 0.05; **: p < 0.01; ***: p < 0.001).
